# A rapid review of the effectiveness, efficiency, and acceptability of surgical hubs in supporting planned care activity

**DOI:** 10.1101/2023.04.20.23288815

**Authors:** Chukwudi Okolie, Jordan Everitt, Alesha Wale, Helen Morgan, Toby Ayres, Hannah Shaw, Ruth Lewis, Alison Cooper, Adrian Edwards

**Author notes:** **Funding statement**: The Evidence Service, Public Health Wales was funded for this work by the Health and Care Research Wales Evidence Centre, itself funded by Health & Care Research Wales on behalf of Welsh Government.

## Abstract

The COVID-19 pandemic further exacerbated disruptions to elective care services in the UK, leading to longer waits for treatment and a growing elective surgery backlog. There have been growing calls for the creation of surgical hubs to help reduce this backlog. Surgical hubs aim to increase surgical capacity by providing quicker access to procedures, as well as facilitate infection control by segregating patients and staff from emergency care. This rapid review aimed to assess the effectiveness, efficiency, and acceptability of surgical hubs in supporting planned care activity, to inform the implementation of these hubs in Wales.

The review identified evidence available up until January 2023. Twelve primary studies were included, eight of which used comparative methods. Most of the studies were conducted during the COVID-19 pandemic and described surgical hubs designed mainly to mitigate the transmission of the severe acute respiratory syndrome coronavirus 2 (SARS-CoV-2). Outcome measures reported included clinical, performance, economic, and patient reported outcomes across a variety of different surgical disciplines. Most of the studies did not describe surgical hubs based on their structure, i.e., standalone, integrated, or ring-fenced hubs.

The evidence relating to the impact of surgical hubs on clinical outcomes appeared to be heterogenous and limited. Included studies did not appear to control for the impact of the COVID-19 pandemic on outcomes. Evidence of the impact of surgical hubs on performance outcomes such as efficiency, utilisation/usage, volume of surgeries/treatments, performance, cancellations, and time from diagnosis to treatment is limited. Evidence relating to the economic impact of surgical hubs is also limited, however there is evidence to suggest that total average costs are lower in surgical hubs when compared to general hospitals. Evidence relating to the impact of surgical hubs on patient reported outcomes is limited but indicates there may be a positive effect on patient satisfaction and compliance.

Considerable variation in the types of surgical hubs reviewed, surgical disciplines, along with the small number of comparative studies, as well as methodological limitations across included studies, could limit the applicability of these findings.

## 1. BACKGROUND

### 1.1 Who is this review for?

This Rapid Review was conducted as part of the Health and Care Research Wales Evidence Centre work programme. The above question was suggested by the Royal College of Surgeons of Edinburgh to inform resilient elective care strategies and to support the implementation of surgical hubs across Wales.

### 1.2 Background and purpose of this review

Over the last decade, the elective care waiting list in the UK has grown substantially. Prior to the COVID-19 pandemic, there were already 4.43 million people waiting for elective care services (British Medical Association, 2023). A combination of reduced NHS funding, staffing, and capacity were likely causes of this disruption in services (Mallorie, 2023). The COVID-19 pandemic significantly added to these disruptions, creating an unprecedented backlog in elective care.

Dealing with the backlog of elective surgical services is a critical concern for the NHS in the UK. The ability to have a resilient elective care system during emergency pressures, such as the yearly winter pressures, is also pressing.

There have been growing calls for separation of elective and emergency care, and the creation of surgical hubs to help deal with the elective care backlog (Royal College of Surgeons of England, 2022). Surgical hubs are a key element of the elective recovery strategy for the NHS and are defined as protected facilities dedicated entirely to elective care, with ring-fenced resources that allow them to stay active even when emergency pressures arise (Briggs et al., 2022). These hubs aim to increase surgical capacity by providing quicker access to procedures, and to facilitate infection control by segregating patients and staff from emergency departments (Royal College of Surgeons of England, 2022).

There are three main types of elective surgical hubs (GIRFT, 2022). These are:

- ‘Stand-alone hubs’ (i.e., elective surgical unit in a dedicated building fully separate from any acute provision)
- ‘Integrated hubs’ (i.e., elective surgical unit within an existing acute hospital site)
- ‘Ring-fenced hubs’ (i.e., elective surgical unit exists as dedicated area within an existing acute hospital)

There are currently 91 operational surgical hubs in England, with over 50 new hubs set to open across the country by 2024 (Department of Health and Social Care, 2022). In Wales, the rollout of surgical hubs has been envisioned in the Welsh Government Planned Care Plan (Welsh Government, 2022). However, little is currently known about their effectiveness.

The purpose of this rapid review is to assess the effectiveness, efficiency, and acceptability of surgical hubs in supporting planned care activity, to inform the implementation of these hubs in Wales. In assessing this, the review will attempt to address the following review sub-uestions:

- What is the effectiveness of surgical hubs in delivering elective care (e.g., treatment numbers, timing, and clinical outcomes), in particular during periods of emergency or high pressure?
- What is the most effective model (stand-alone, integrated, or ring-fenced) for surgical hubs in maintaining resources and to stay active when emergency pressures rise?
- What is the most effective work force model for surgical hubs?
- How far are people willing to travel/what is the travel experience?
- What innovative roles have been developed to deal with any workforce challenges?
- In the UK, have any contract/practice adaptions been made to attract staff e.g., travel time, training opportunities, mixed model of private and NHS staff?
- What is the most effective governance model for commissioning and running a surgical Hub, in particular when covering populations outside of one or more health organisation boundaries and when cross-organisational working is required?
- Which speciality or for which procedure is the surgical hub model most effective and efficient?
- What are patients views of surgical hubs (pros and cons)?
- Have surgical hubs been able to recruit to full headcount and what roles have proved more challenging?
- How far are staff willing to travel?

## 2. RESULTS

### 2.1 Overview of the Evidence Base

Twelve primary studies were eligible for inclusion in this rapid review (six quasi-experimental, two cohort and four case series). Eight studies used comparative research methods, i.e., compared surgical hubs with other surgical units or compared the period a surgical hub was in operation with the period predating the establishment of the surgical hub, while five studies were non-comparative, i.e., described a single centre.

Included studies were conducted in the UK (n = 7), the Netherlands (n = 3), and Italy (n = 2). Two studies each were focussed on orthopaedic surgery, colorectal cancer surgery, and ophthalmic surgery, while one study each focussed on gynaecological oncology surgery, head and neck cancer surgery, and robotic surgery for colorectal and urological cancer. Three studies were focussed on multiple surgical specialities as opposed to a single specialty. A detailed summary of included studies can be found in Table 1.

**Table 1:**
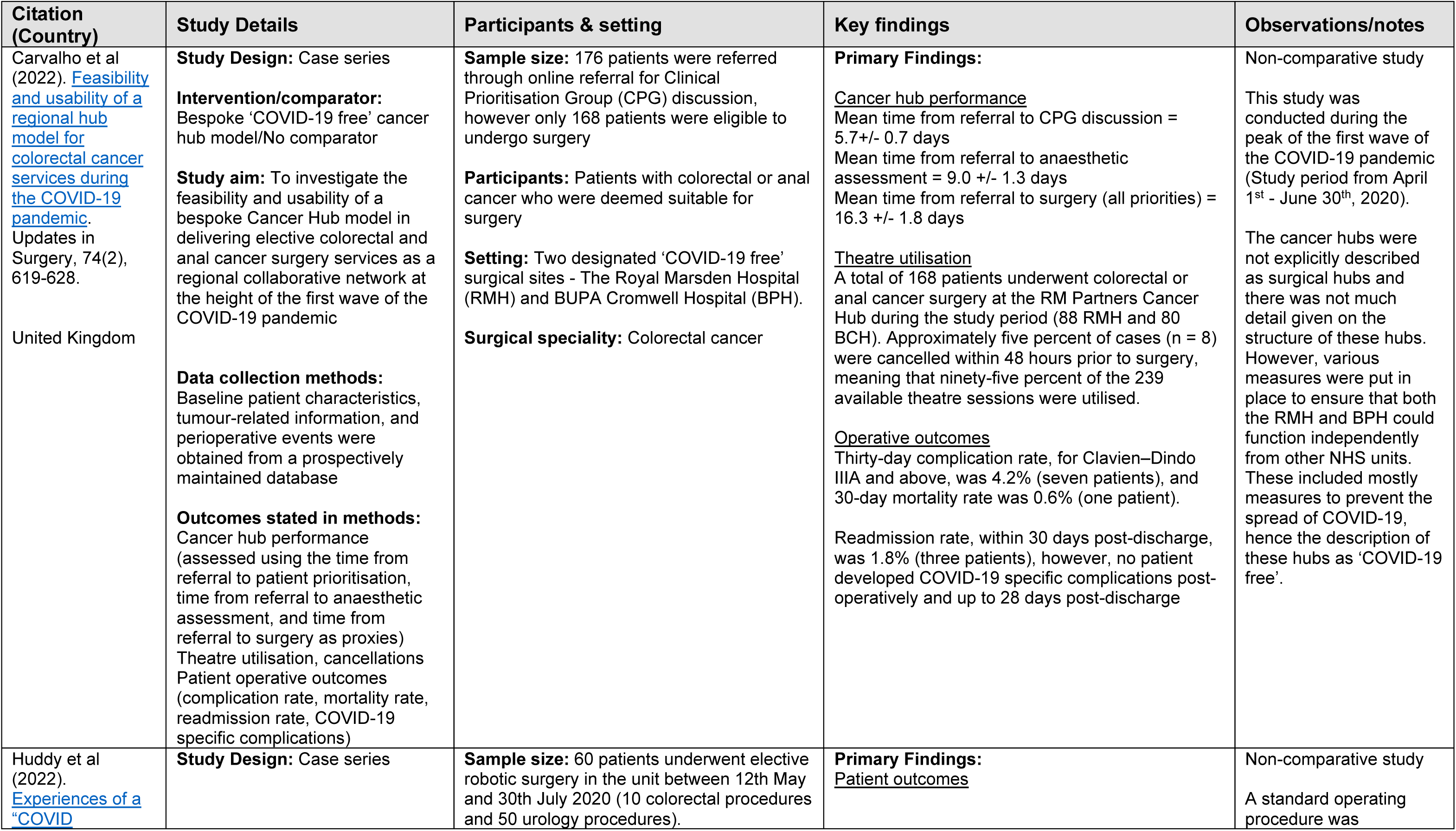
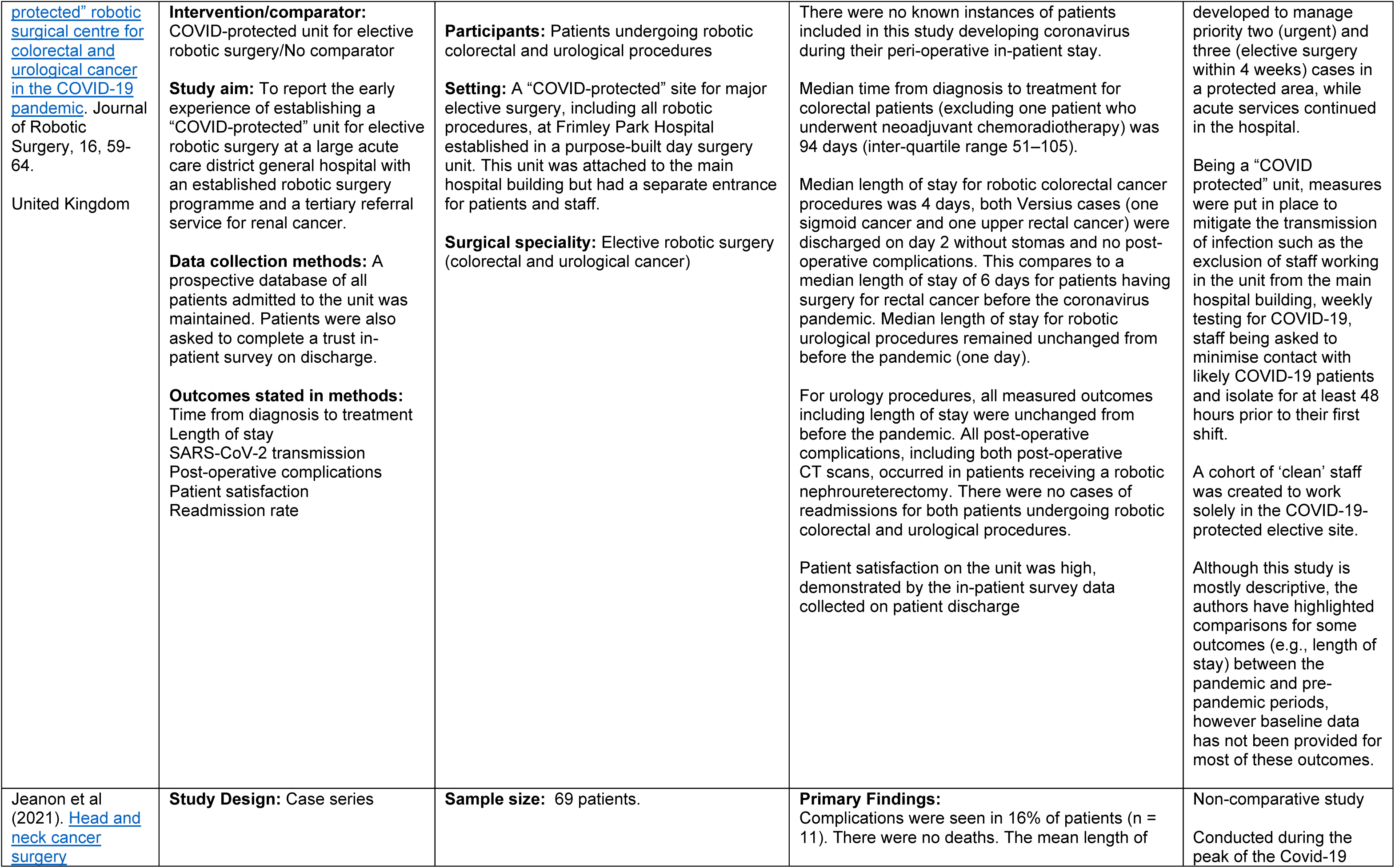
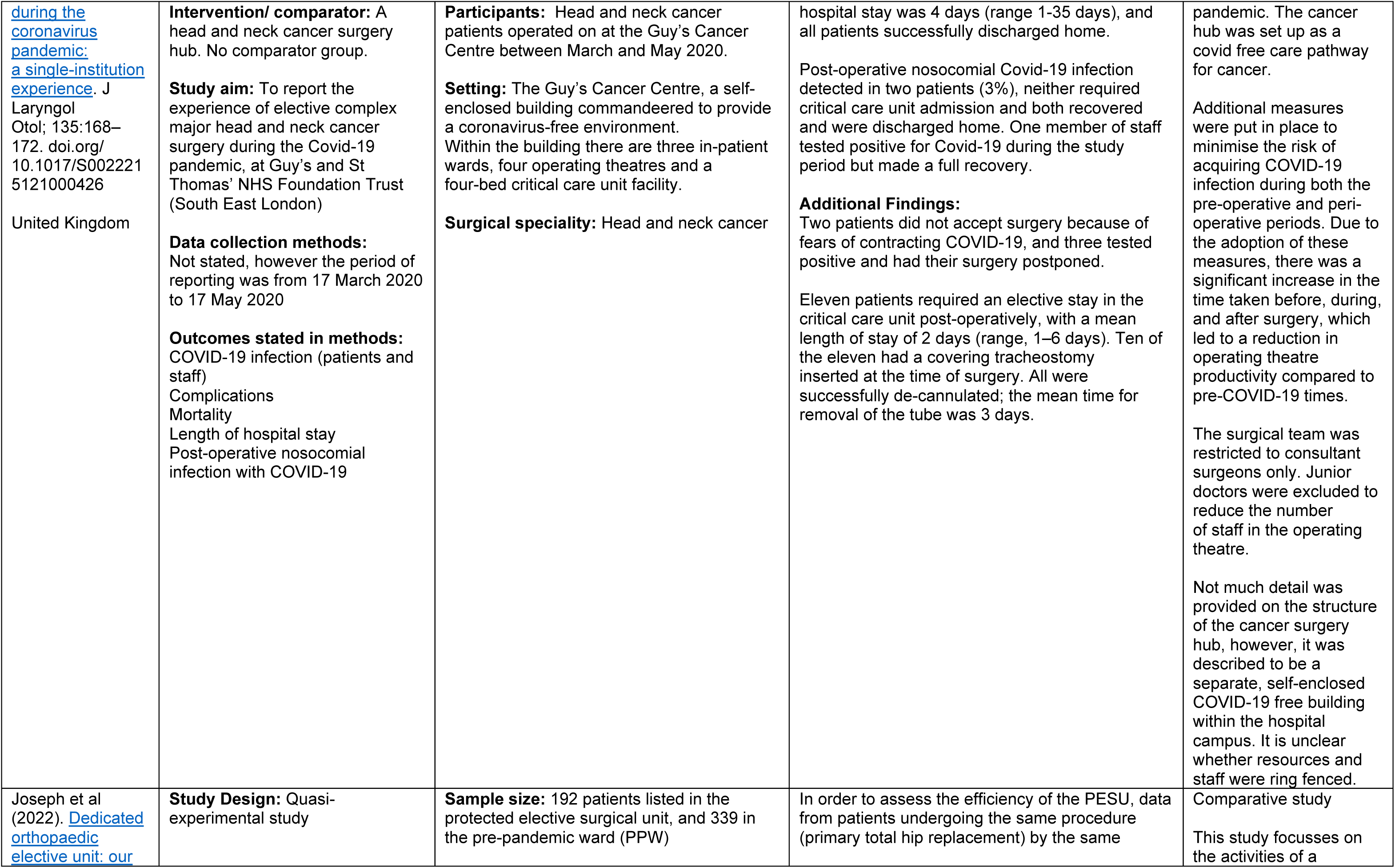
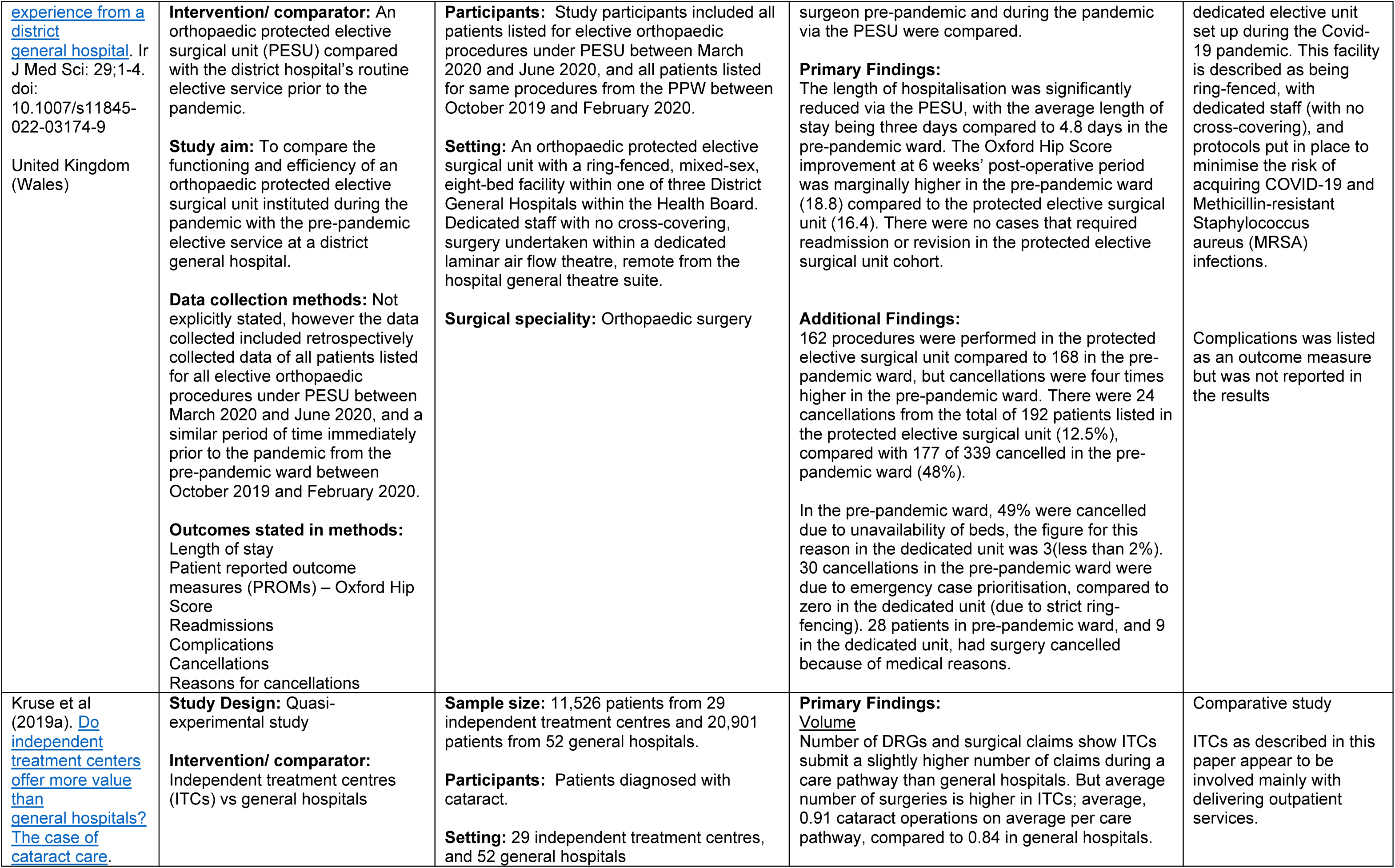
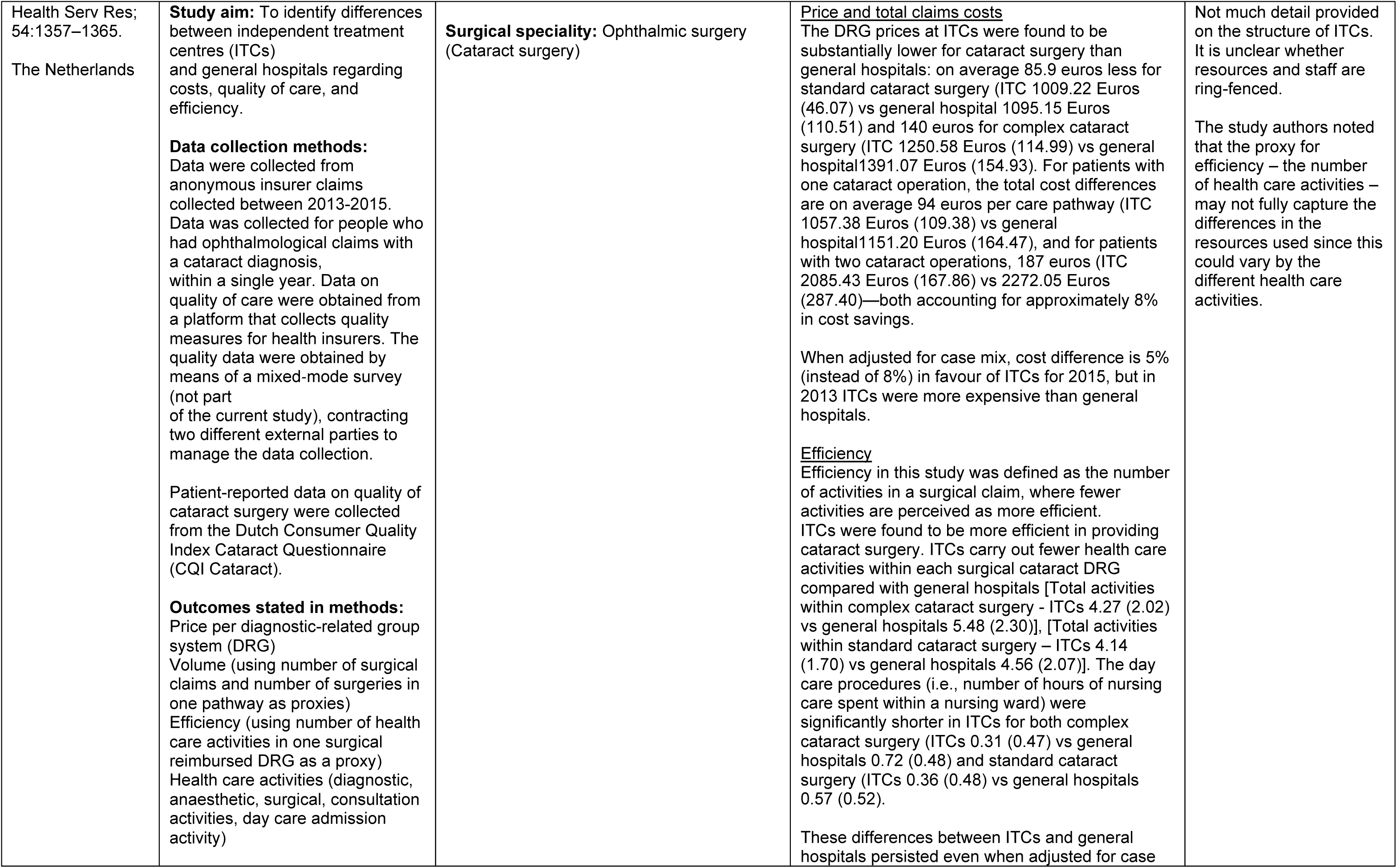
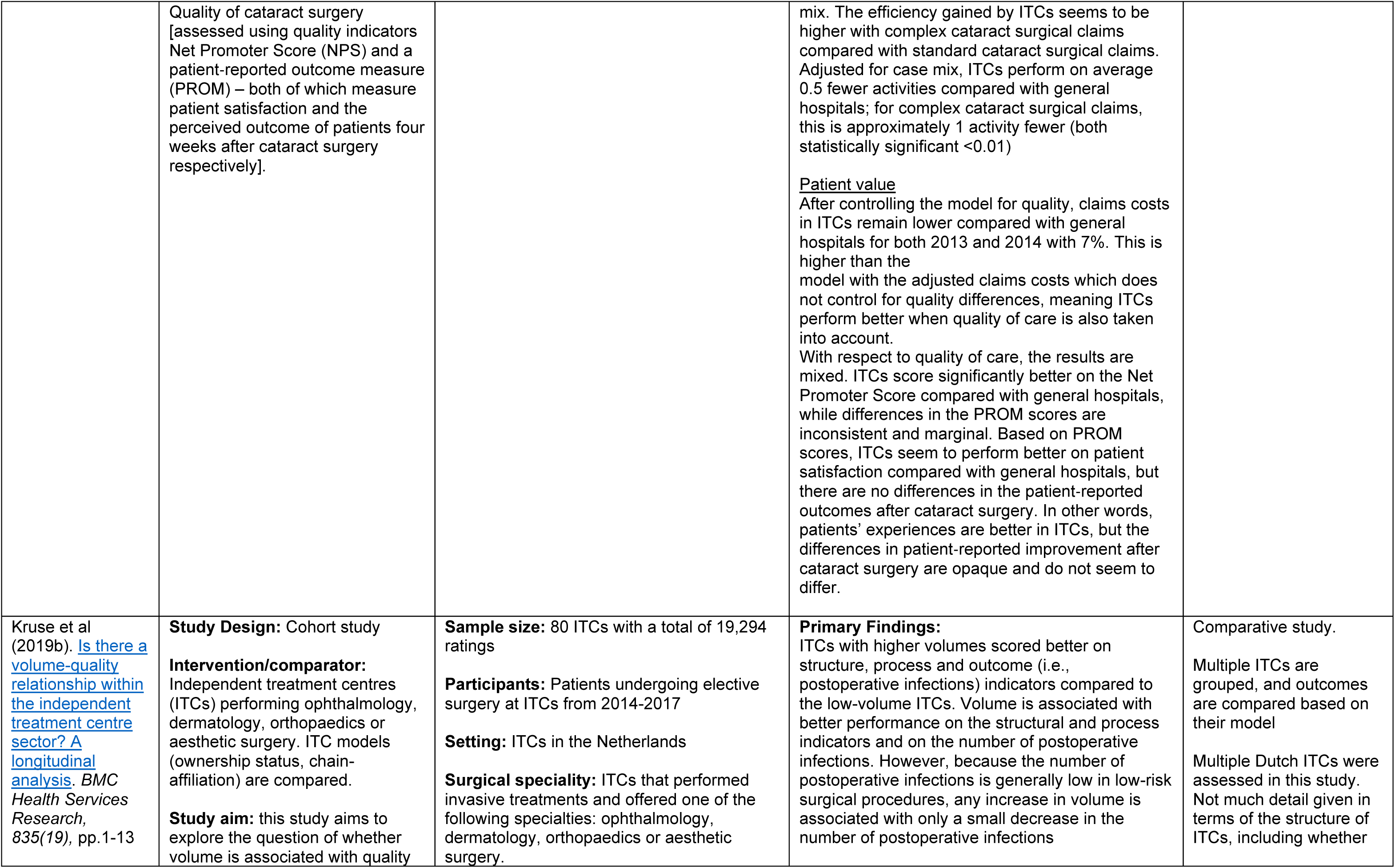
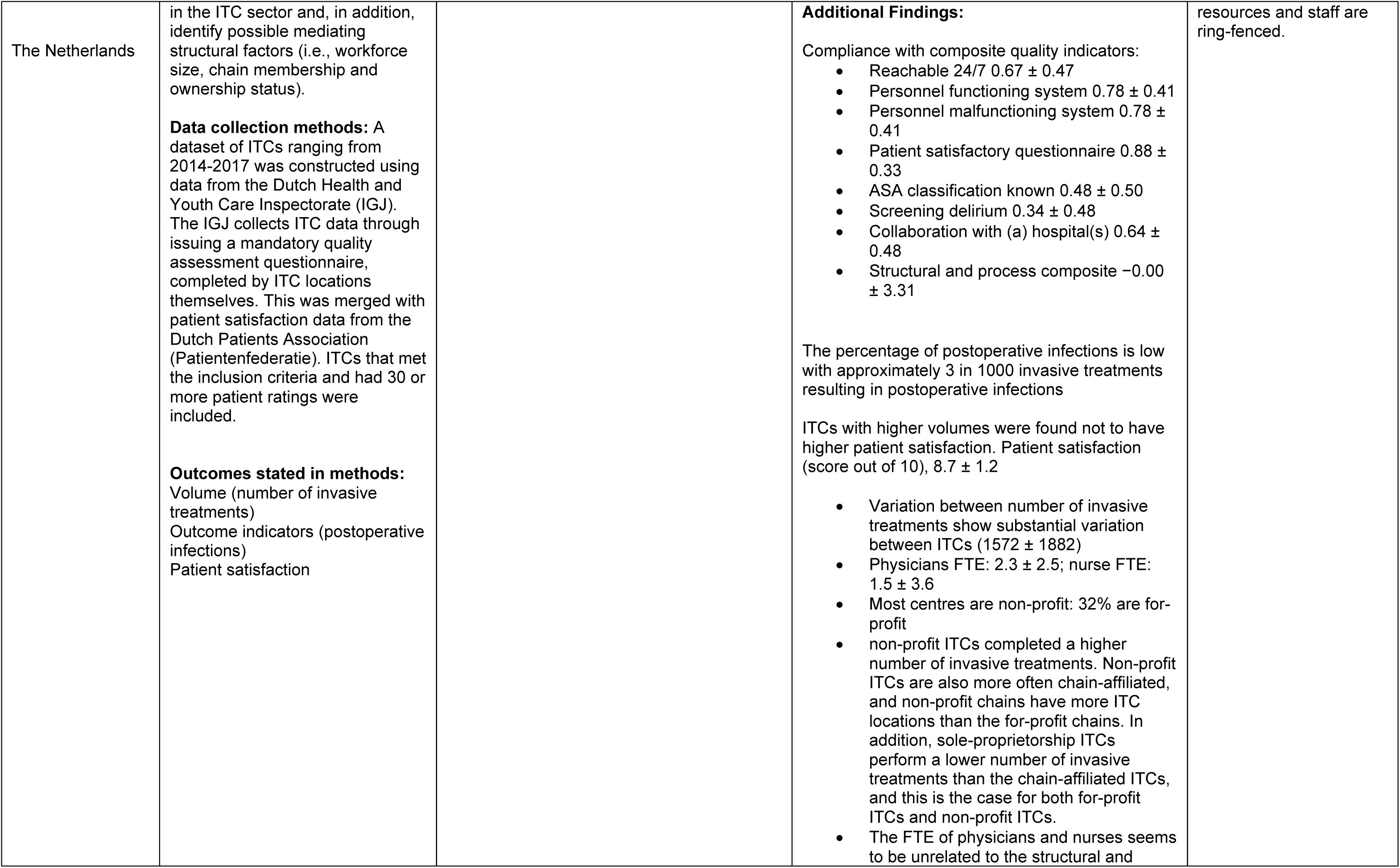
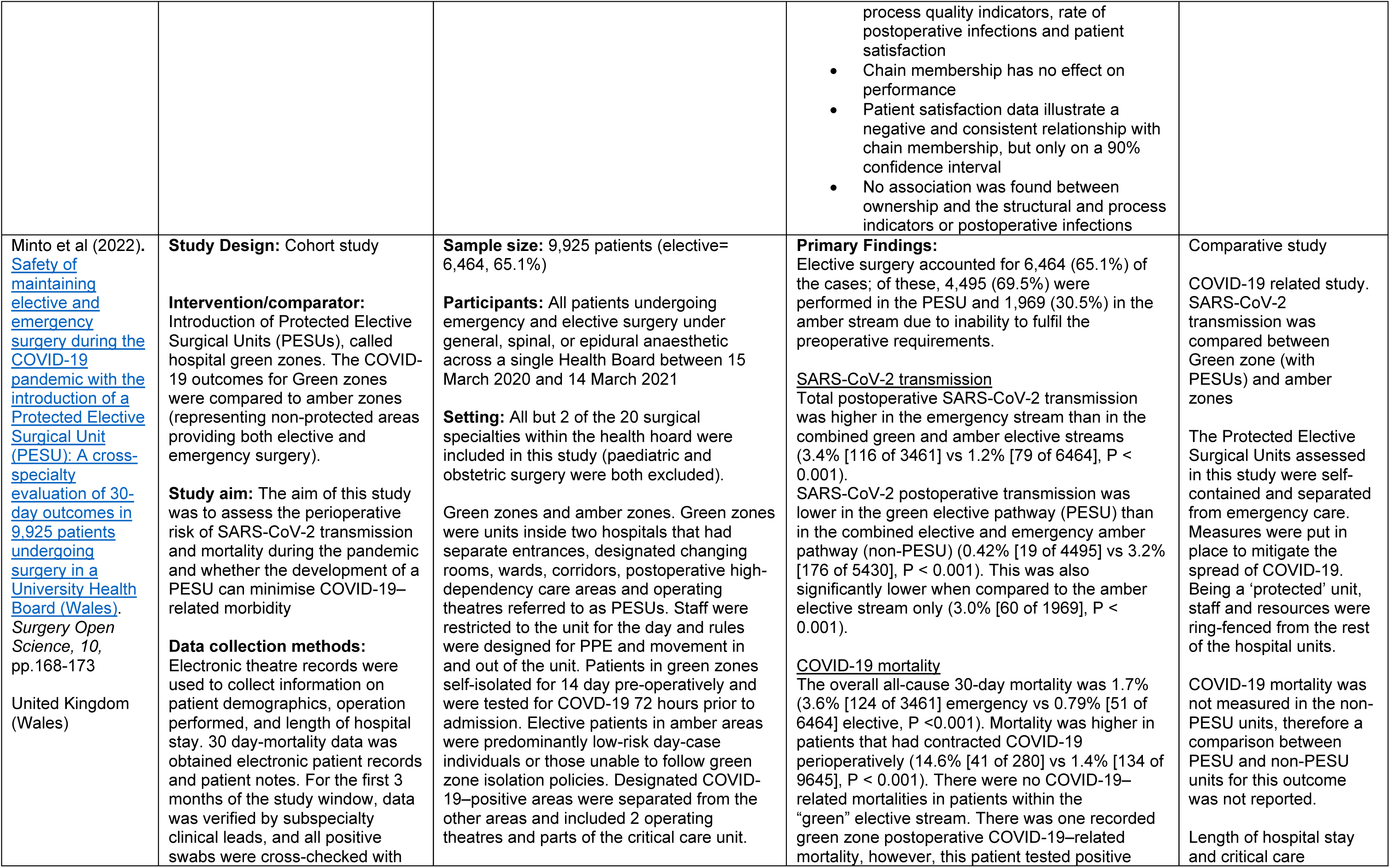
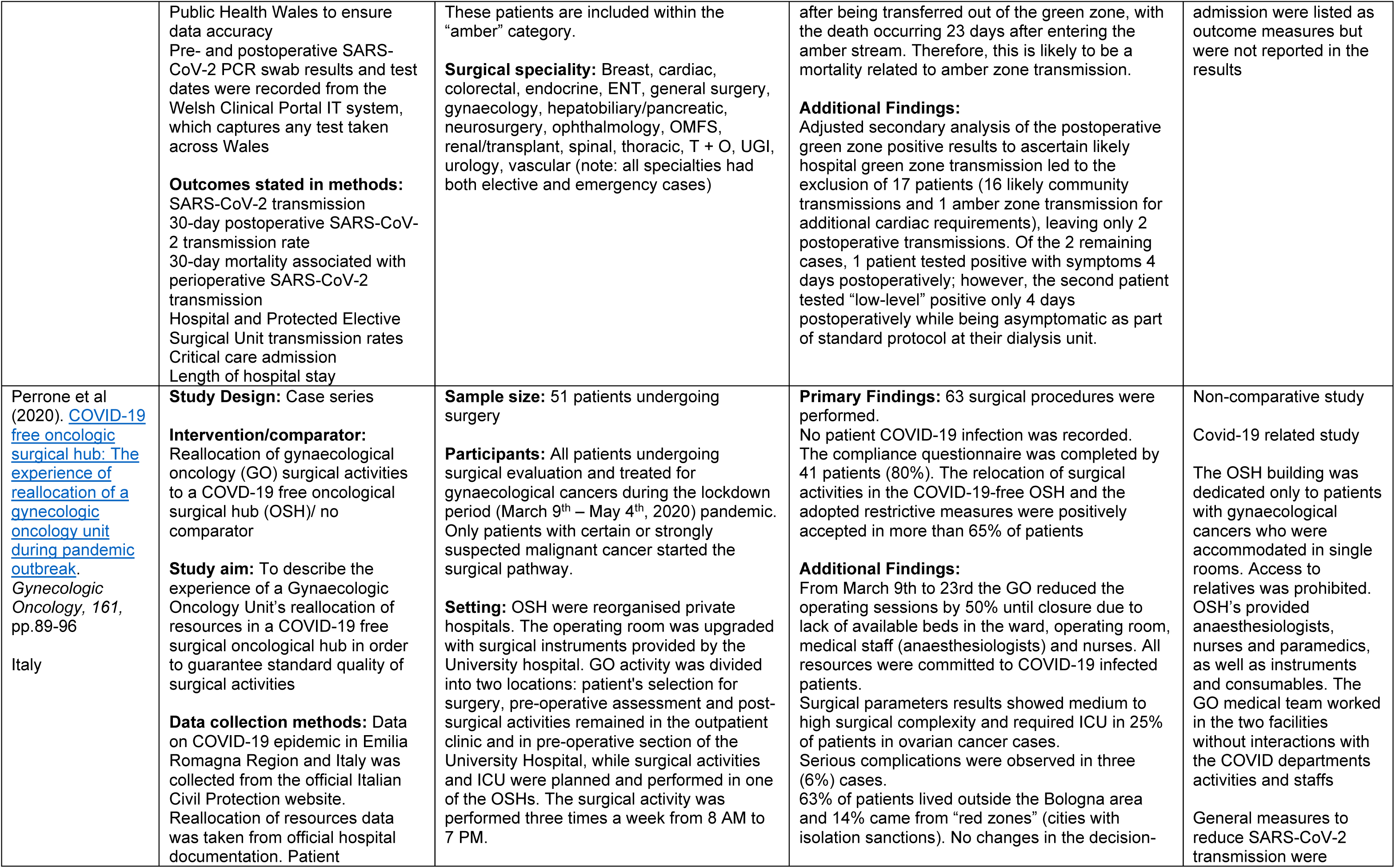
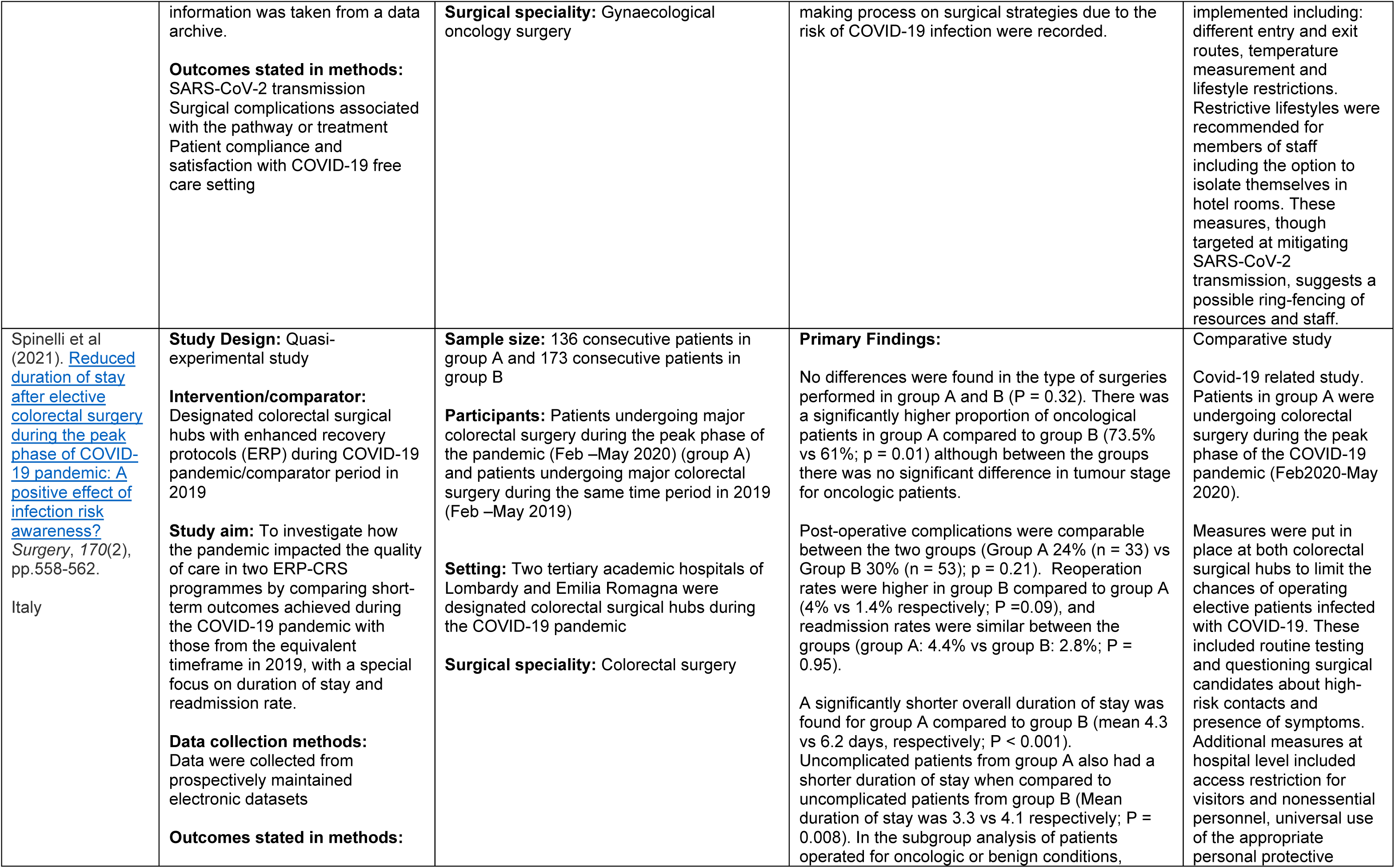
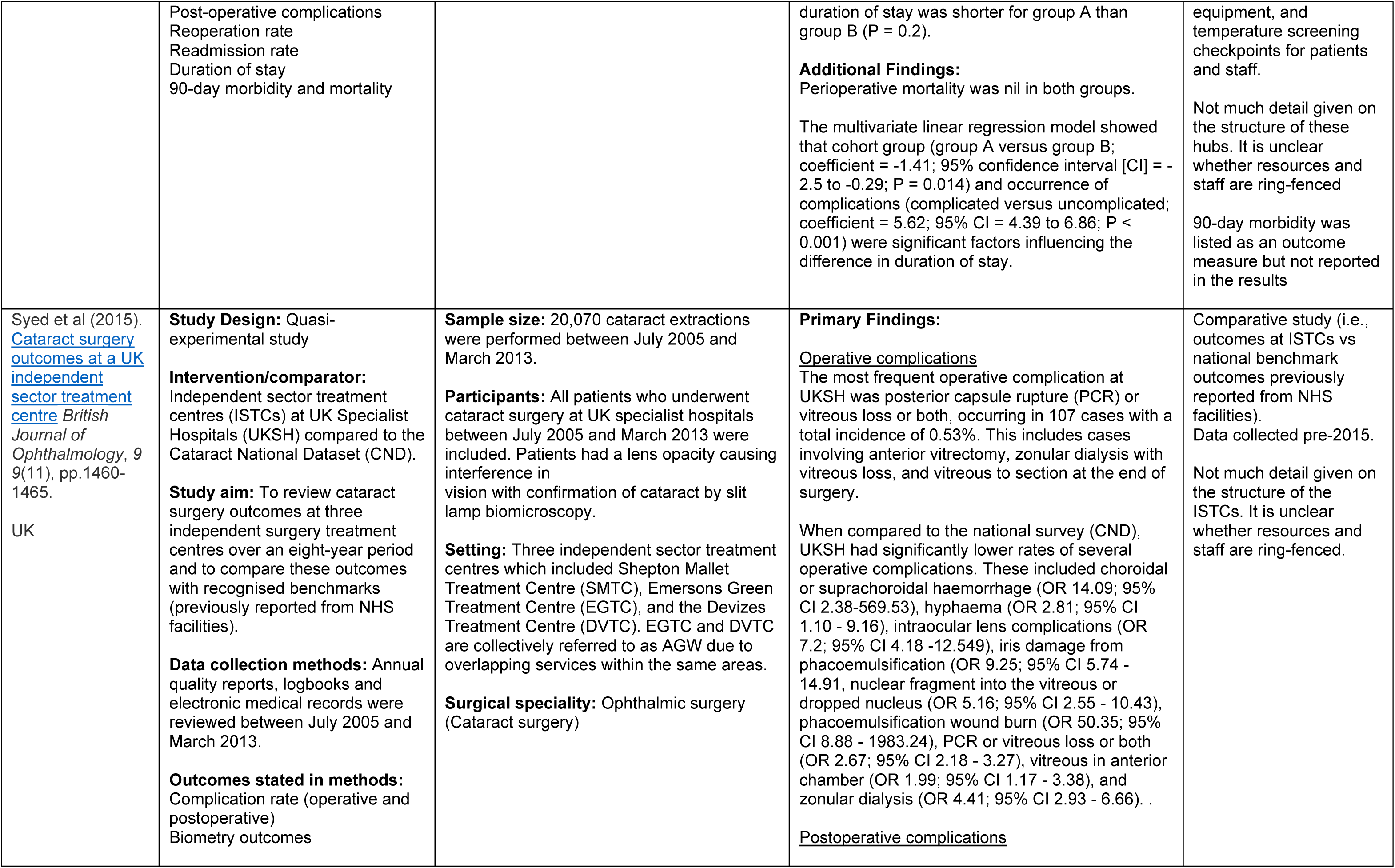
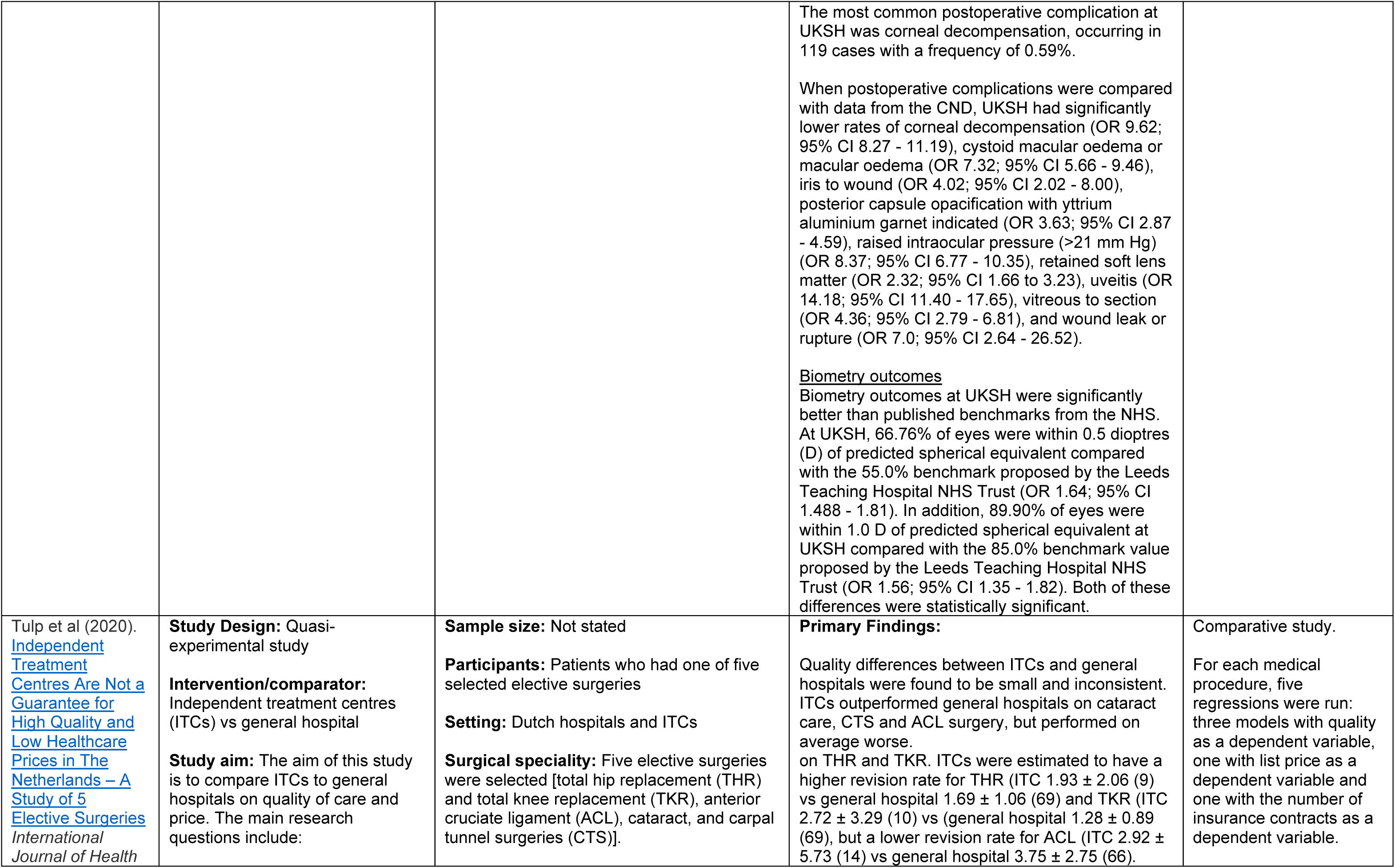
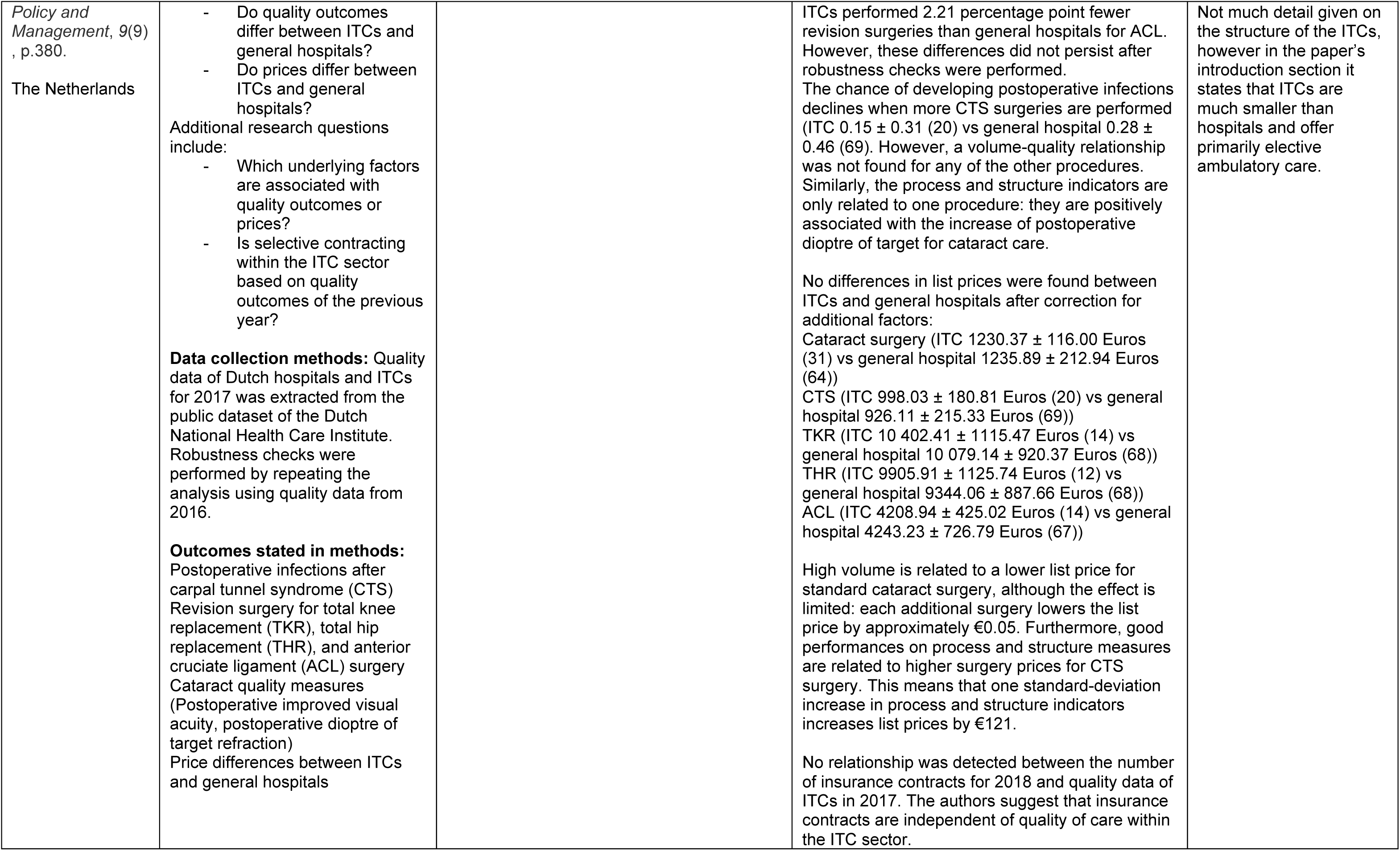
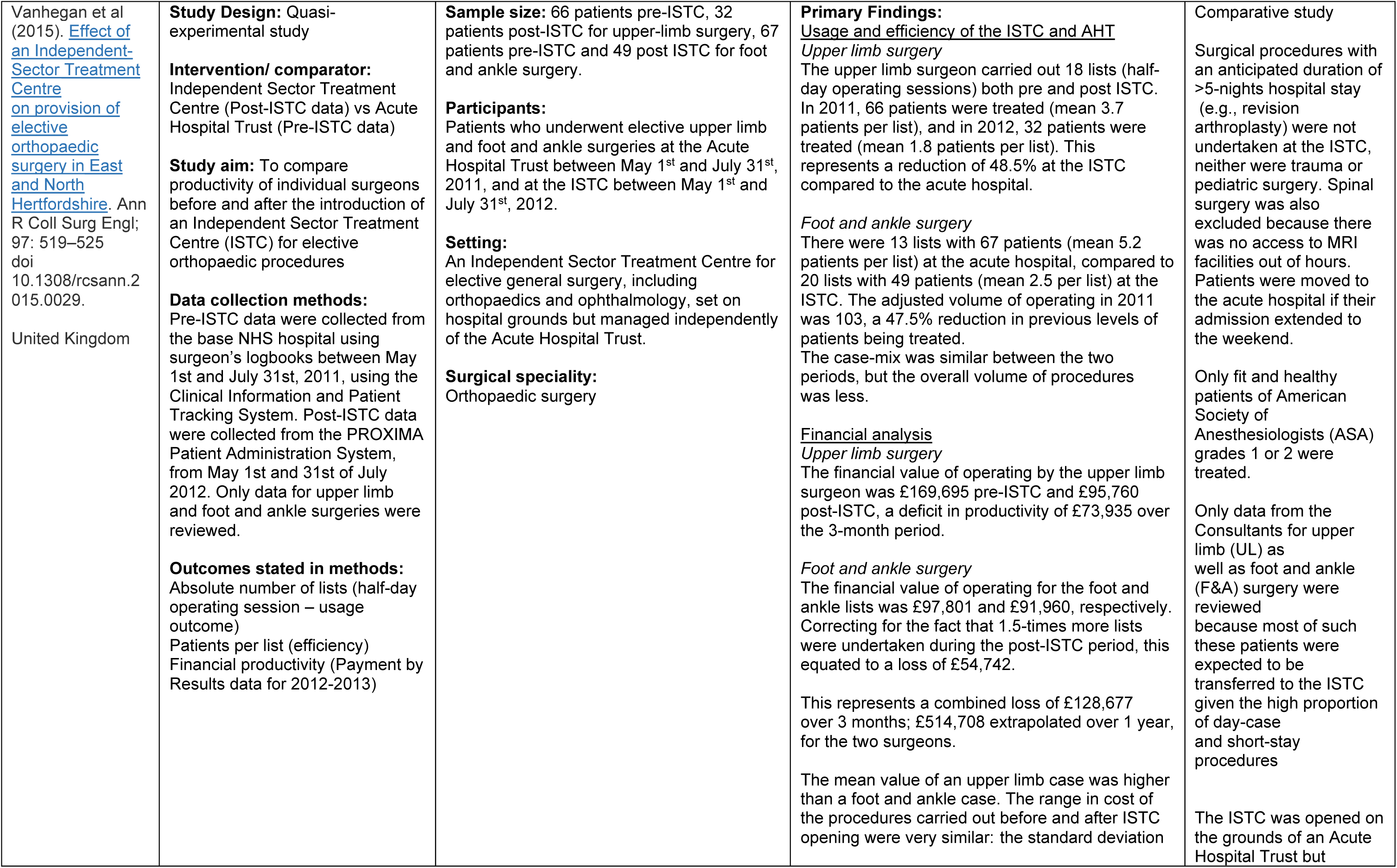
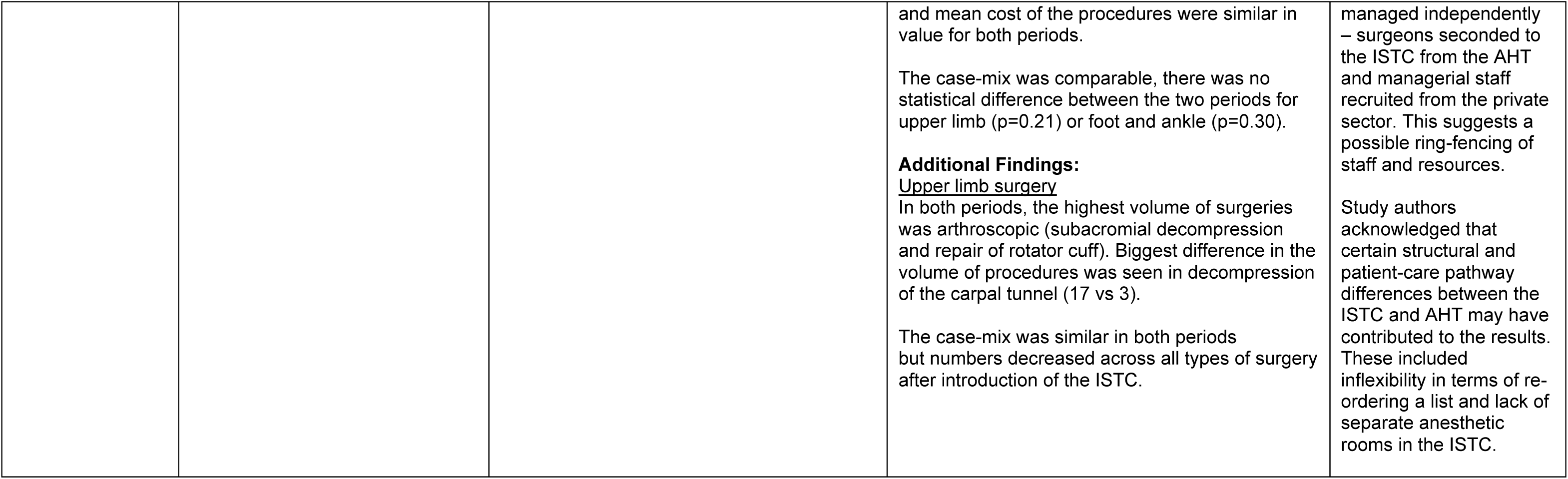
Summary of included studies

The majority of included studies were conducted during the COVID-19 pandemic (n = 7) and reported on surgical hubs designed mainly to mitigate the transmission of SARS-CoV-2. As a result, COVID-19 free hubs or COVID-19 protected sites were the most common surgical hubs reported in the included studies (n = 5). Other surgical hubs included independent treatment centres (ITC) (n = 3), independent sector treatment centres (ISTC) (n = 2) and protected elective surgical units (PESU) (n = 2). Most of the included studies did not describe surgical hubs based on their structure, i.e., standalone, integrated, or ring-fenced hubs. Only two studies (Joseph et al., 2022, Minto et al., 2022) explicitly described surgical facilities with ring-fenced or dedicated capacity/resources. Joseph et al (2022) explored outcomes at a dedicated orthopaedic PESU with ring-fenced resources, embedded within a district general hospital. Minto et al (2022) also evaluated a PESU which was self-contained, separated from emergency care, and had staff and resources ring-fenced from other hospital units. Due to the heterogeneity of surgical facilities included in this review, a narrative synthesis approach was used to analyse data and present findings.

The methodological quality of included studies was assessed using the appropriate Joanna Briggs Institute (JBI) critical appraisal tool (for quasi-experimental studies, cohort studies and case series). Quality appraisal identified all studies had some methodological limitations. Further details of the quality appraisal can be found in section 6.

For this review, only comparative studies comparing surgical hubs with other surgical units or comparing time periods before and after establishment of a surgical hub, were analysed when evaluating effectiveness outcomes, as these studies are better placed to determine cause and effect relationships (Joseph et al 2022, Kruse et al 2019a, Kruse et al 2019b, Minto et al 2022, Spinelli et al 2021, Syed et al 2015, Tulp et al 2020 Vanhegan et al 2015). For patient-reported outcomes such as patient satisfaction and compliance, data from both comparative and non-comparative studies were utilised (Huddy et al 2022, Joseph et al 2022. Kruse et al 2019a, Kruse et al 2019b, Perrone et al 2020).

Figure 1 outlines the outcome measures reported in all 12 included primary studies. The primary studies have been split into comparative and non-comparative and the outcomes categorised as clinical, performance, economic and patient-reported outcomes.

**Figure 1.**
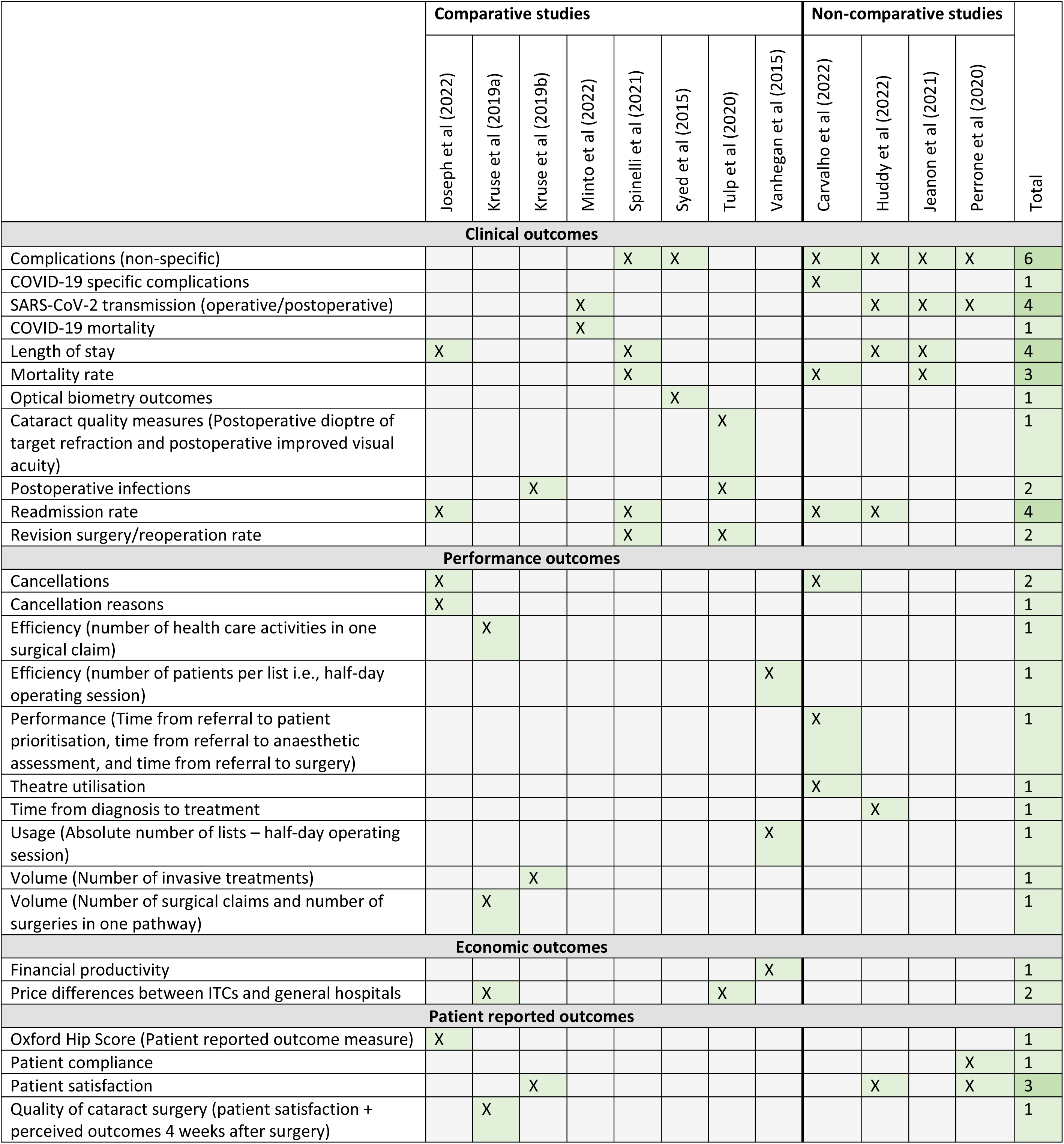
Outcomes map

### 2.2 Impact of surgical hubs on clinical outcome measures

Ten studies (six comparative and four non-comparative) reported a range of clinical outcomes. These included length of hospital stay, mortality, complications (non-specific and COVID-19 related), SARS-CoV-2 transmission, COVID-19 mortality, clinical outcome measures for cataract surgery (postoperative dioptre of target refraction and postoperative improved visual acuity), postoperative infections, readmissions, and reoperation rate. Only findings from comparative studies are reported here. The results of the non-comparative studies are provided in Table 1.

#### Length of hospital stay

Length of hospital stay was reported in two studies which compared outcomes between patients undergoing surgery at surgical hub facilities during the COVID-19 pandemic with patients who had undergone surgery pre-pandemic.

Joseph et al (2022) compared the functioning and efficiency of an orthopaedic PESU instituted during the COVID-19 pandemic with the pre-pandemic elective service at a general hospital ward. The **length of hospitalisation was significantly reduced at the PESU compared to the pre-pandemic ward** (mean length of hospitalisation 3 days vs 4.8 days). However, this study did not appear to control for the impact of the pandemic in trying to speed up hospital discharge.

Spinelli et al (2021) compared outcomes of patients undergoing major colorectal surgery with an enhanced recovery protocol during the COVID-19 pandemic (group A) with those from an equivalent timeframe before the pandemic in 2019 (group B). This study found a **significantly shorter overall duration of stay for group A patients compared to group B** (mean duration of stay 4.3 days vs 6.2 days; p < 0.001). Uncomplicated patients from group A also had a shorter duration of stay when compared to uncomplicated patients from group B (Mean duration of stay 3.3 days vs 4.1 days; p = 0.008). This study did not control for the impact of the pandemic on outcomes.

#### Readmission rates

Only one study provided data comparing readmission rates in surgical hub patients during the COVID-19 pandemic with patients who had undergone surgery pre-pandemic.

Spinelli et al (2021) found that readmission rates were similar between surgical hub patients undergoing surgery with enhanced recovery protocols during the peak phase of pandemic, compared to patients who had undergone surgery before the pandemic (n = 6 [4.4%] vs n = 5 [2.8%]; p = 0.95).

#### Reoperation rate/revision surgery

Reoperation rates or revision surgeries were reported in two comparative studies.

Tulp et al (2020) compared quality of care and price differences between Dutch ITCs and general hospitals. This study found that ITCs have a higher revision rate (within one year) for total hip replacement (THR) (1.93% ± 2.06 vs 1.69% ± 1.06) and total knee replacement (TKR) (2.72% ± 3.29 vs 1.28% ± 0.89), but a lower revision rate for anterior cruciate ligament (ACL) surgery (2.92% ± 5.73 vs 3.75% ± 2.75), when compared with general hospitals. However, these differences did not persist after robustness checks (use of quality data from 2016, exclusion of specialist and academic hospitals, and inclusion of outliers) were performed.

Spinelli et al (2021) found that reoperation rates were higher in surgical hub patients undergoing surgery with enhanced recovery protocols during the peak phase of pandemic, compared to patients who had undergone surgery before the pandemic (4% vs 1.4%; p = 0.09). This study did not control for the impact of the pandemic on outcomes.

#### Complications (non-specific operative/post-operative)

Surgical complications were reported in two comparative studies.

Syed et al (2015) reviewed cataract surgery outcomes at three ISTCs established by the UK Specialist Hospitals (UKSH) and compared these with recognised benchmarks previously reported from NHS facilities. When compared to published benchmarks from the Cataract National Dataset (CND), UKSH had **significantly lower rates of several operative complications**. These included choroidal or suprachoroidal haemorrhage (odds ratio [OR] 14.09; 95% confidence interval [CI] 2.38 – 569.53; p<0.05), hyphaema (OR 2.81; 95% CI 1.10 – 9.16; p<0.05), intraocular lens complications (OR 7.2; 95% CI 4.18 – 12.549; p<0.05), iris damage from phacoemulsification (OR 9.25; 95% CI 5.74 – 14.91; p<0.05), nuclear fragment into the vitreous or dropped nucleus (OR 5.16; 95% CI 2.55 – 10.43; p<0.05), phacoemulsification wound burn (OR 50.35; 95% CI 8.88 – 1983.24; p<0.05), PCR or vitreous loss or both (OR 2.67; 95% CI 2.18 – 3.27; p<0.05), vitreous in anterior chamber (OR 1.99; 95% CI 1.17 – 3.38; p<0.05), and zonular dialysis (OR 4.41; 95% CI 2.93 – 6.66; p<0.05). Similarly, when postoperative complications were compared with data from the CND, UKSH had **significantly lower rates** of corneal decompensation (OR 9.62; 95% CI 8.27 – 11.19; p<0.05), cystoid macular oedema or macular oedema (OR 7.32; 95% CI 5.66– 9.46; p<0.05), iris to wound (OR 4.02; 95% CI 2.0– 8.00; p<0.05), posterior capsule opacification with yttrium aluminium garnet indicated (OR 3.63; 95% CI 2.87 – 4.59; p<0.05), raised intraocular pressure (>21 mm Hg) (OR 8.37; 95% CI 6.77 – 10.35; p<0.05), retained soft lens matter (OR 2.32; 95% CI 1.66 to 3.23; p<0.05), uveitis (OR 14.18; 95% CI 11.40 – 17.65; p<0.05), vitreous to section (OR 4.36; 95% CI 2.79 – 6.81; p<0.05), and wound leak or rupture (OR 7.0; 95% CI 2.64 – 26.52; p<0.05).

Spinelli et al (2021) found that post-operative complication rates were comparable between surgical hub patients undergoing surgery with enhanced recovery protocols and patients who had undergone surgery before the pandemic [24% (n = 33) vs 30% (n = 53); p = 0.21].

#### COVID-19-related outcomes

A single comparative study, from the UK, reported SARS-CoV-2 transmission rates at a surgical hub facility.

Minto et al (2022) assessed whether the development of a PESU can minimise SARS-CoV-2 transmission and mortality. The results showed that **SARS-CoV-2 postoperative transmission was significantly lower in the PESU than in the non-PESU** facility (0.42% vs 3.2% p < 0.001). COVID-19 mortality was not measured in the non-PESU units, therefore a comparison between PESU and non-PESU units for this outcome was not reported.

#### Optical biometry outcomes

One UK comparative study reported optical biometry outcomes related to cataract surgery.

Syed et al (2015) found that biometry outcomes at ISTCs established by the UK Specialist Hospitals (UKSH) **were significantly better than published benchmarks previously reported from NHS facilities**. At UKSH, 66.76% of eyes were within 0.5 dioptres (D) of predicted spherical equivalent compared with the 55.0% benchmark proposed by the Leeds Teaching Hospital NHS Trust (OR 1.64; 95% CI 1.488 – 1.81; p<0.01). In addition, 89.90% of eyes were within 1.0 D of predicted spherical equivalent at UKSH compared with the 85.0% benchmark value proposed by the Leeds Teaching Hospital NHS Trust (OR 1.56; 95% CI 1.35 – 1.82; p<0.01). Both differences were statistically significant.

#### Cataract quality measures

One Dutch comparative study reported quality measures for cataract surgery (postoperative improved visual acuity and postoperative dioptre of target refraction).

Tulp et al (2020) compared quality differences between Dutch ITCs and general hospitals and found that ITCs outperformed general hospitals on postoperative improved visual acuity (85.58% ± 9.81 vs 83.10% ± 7.25) and postoperative dioptre of target refraction (94.87% ± 3.32 vs 93.78% ± 3.45), however differences were small.

#### Postoperative infections

One Dutch study made comparisons on postoperative infections between a surgical hub facility and other clinical settings.

Tulp et al (2020) found that the chance of developing postoperative infections within 30 days of carpal tunnel syndrome surgery was less at ITCs compared to general hospitals (0.15% ± 0.31 vs 0.28% ± 0.46).

#### 2.2.1 Bottom line results for the impact of surgical hubs on clinical outcome measures

There is evidence to suggest that surgical hubs can be effective at improving clinical outcomes such as length of hospital stay, operative and post-operative complications, and cataract surgery quality measures in certain surgical fields. However, the evidence supporting the use of these facilities in reducing readmission rates appears to be limited as each of these outcomes were often only reported by a single comparative study. Included studies did not control for the impact of the pandemic on outcomes.

### 2.3 Impact of surgical hubs on performance outcome measures

Six studies (Four comparative and two non-comparative) reported a range of performance outcomes. These included efficiency, utilisation/usage, volume of surgeries/treatments, performance, cancellations, and time from diagnosis to treatment. Only findings from comparative studies are reported here. The results of the non-comparative studies are provided in Table 1.

### Efficiency

Two comparative studies (one from the Netherlands and the other from the UK) reported efficiency measures.

Kruse et al (2019a) focussed on cataract care and sought to identify differences between Dutch independent treatment centres (ITCs) and general hospitals regarding costs, quality of care, and efficiency. Efficiency in this study was defined as the number of activities in a surgical claim, where fewer activities are perceived as more efficient. ITCs were found to be more efficient than general hospitals in providing cataract surgery, i.e., ITCs carried out fewer health care activities within each surgical cataract claim compared to general hospitals – [Total activities within complex cataract surgery – ITCs 4.27 (2.02) vs general hospitals 5.48 (2.30)], [Total activities within standard cataract surgery – ITCs 4.14 (1.70) vs general hospitals 4.56 (2.07)]. Differences persisted even when adjusted for case mix.

Vanhegan et al (2015) investigated the effect on productivity of operating theatres working in an independent sector treatment centre (ISTC) compared with those working in the Acute Hospital Trust (AHT). Efficiency in this study was assessed in terms of the number of patients per surgeon list – with a ‘list’ defined as a half-day operating session. The implementation of the ISTC was found to be detrimental to departmental efficiency, with <50% of the number of patients being treated. For upper limb surgery, a mean of 3.7 patients per list were treated at the AHT (pre-ISTC) compared to a mean of 1.8 patients per list treated at the ISTC. This represented a reduction of 48.5% at the ISTC compared to the acute hospital. For foot and ankle surgery, a mean of 5.2 patients per list were treated at the acute hospital, compared to a mean of 2.5 patients per list at the ISTC. This represented a 47.5% reduction in previous levels of patients being treated.

### Utilisation/Usage

One study (from the UK) assessed the performance of surgical hubs by reporting on their utilisation or usage and made comparisons between surgical hub facilities and other clinical settings.

Vanhegan et al (2015) used the absolute number of lists as a reflection of a consultants’ timetabled use of the ISTC, and found that for upper limb surgery, there were 18 lists with 66 patients pre-ISTC and 18 lists with 32 patients post-ISTC. The case-mix was similar in both periods, but numbers decreased across all types of upper limb and foot and ankle surgeries for work done after introduction of the ISTC. For foot and ankle surgery, there were 13 lists with 67 patients pre-ISTC and 20 lists with 49 patients post-ISTC. The case-mix between the two periods was found to be comparable, but the overall volume of procedures was less.

### Volume

Two comparative studies reported the volume of surgeries or treatments as an assessment of a surgical hub’s performance.

Kruse et al (2019a) used the total number of surgeries and surgical claims during a care pathway as a proxy for volume. ITCs were found to submit a slightly higher number of claims during a care pathway than general hospitals (Mean) 1.45 (SD 0.63) vs 1.41 (SD 0.63). The average number of surgeries was also found to be higher in ITCs with a mean of 0.91 (SD 0.81) cataract operations on average per care pathway, compared to a mean of 0.84 (SD 0.77) in general hospitals.

Kruse et al (2019b) used the number of invasive treatments as a proxy for volume and investigated how low-volume and high-volume ITCs performed relative to each other. ITCs with higher volumes were found to score better on structure, process and outcome indicators compared to low-volume ITCs (Mean) 502.40 (SD 1269.82).

### Cancellation of procedures

One comparative study (from the UK) reported comparative data on the cancellation of procedures as an assessment of a surgical hub’s performance.

Joseph et al (2022) compared outcomes between an orthopaedic PESU instituted during the COVID-19 pandemic and a pre-pandemic elective service at a general hospital ward. The study found that cancellations were four times higher in the pre-pandemic ward. The **PESU had a significantly better conversion rate with only 12.5% being cancelled**, compared with 48% of procedures cancelled at the pre-pandemic ward. In the pre-pandemic ward, 49% of procedures (n = 87) were cancelled due to unavailability of beds compared to less than 2% (n = 3) in the PESU for this reason. Thirty cancellations in the pre-pandemic ward were due to emergency case prioritisation compared to zero in the PESU (due to strict ring-fencing). Twenty-eight patients in pre-pandemic ward, and nine in the dedicated unit, had surgery cancelled because of medical reasons.

### 2.3.1 Bottom line results for the impact of surgical hubs on performance outcome measures

Evidence on the impact of surgical hubs on performance outcomes such as efficiency, utilisation/usage, volume of surgeries/treatments, performance, cancellations, and time from diagnosis to treatment is limited. Evidence in relation to the efficiency of these hubs is inconsistent and from a range of surgical disciplines. There is evidence to suggest that surgical hubs are effective at reducing the surgical cancellations, however, this outcome was reported by a single comparative study and as such firm conclusions cannot be made.

## 2.4 Economic outcomes associated with surgical hubs

Three studies (all comparative) reported economic outcome measures. These outcomes included cost differences, total costs and financial productivity.

### Cost differences

Two Dutch studies compared cost differences between ITCs and general hospitals.

Kruse et al (2019a) found that the price per diagnostic-related group (DRG) care product were substantially lower at ITCs than general hospitals for standard cataract surgery (1009.22 Euros (SD 46.07) vs 1095.15 Euros (SD 110.51) and complex cataract surgery (1250.58 Euros (SD 114.99) vs 1391.07 Euros (SD 154.93) using 2015 data. Similarly, total costs were lower at ITCs than general hospitals for patients with one cataract operation (1057.38 Euros (SD 109.38) vs 1151.20 Euros (SD 164.47) and for patients with two cataract operations (2085.43 Euros (SD 167.86) vs 2272.05 Euros (SD 287.40) – both accounting for approximately 8% in cost savings. These differences persisted even after adjusting for case mix.

Tulp et al (2020) found no differences in list price surgery between ITCs and general hospitals after correction for additional factors: Cataract surgery (1230.37 ± 116.00 Euros vs 1235.89 ± 212.94 Euros); carpal tunnel syndrome (998.03 ± 180.81 Euros vs 926.11 ± 215.33 Euros); total knee replacement (10 402.41 ± 1115.47 Euros vs 10 079.14 ± 920.37 Euros); total hip replacement (9905.91 ± 1125.74 Euros vs 9344.06 ± 887.66 Euros); anterior cruciate ligament (4208.94 ± 425.02 Euros vs 4243.23 ± 726.79 Euros). List prices of the first quarter of 2017 were used in the analyses.

### Financial productivity

One comparative UK study reported on financial productivity.

Vanhegan et al (2015) compared productivity outcomes before and after the establishment of an independent sector treatment centre (ISTC) and found that the financial value of operating for upper limb surgery was higher pre-ISTC compared to post-ISTC (£169,695 vs £95,760; p = 0.21). Similar findings were reported for foot and ankle surgery (£97,801 vs £91,960; p = 0.30). The operative case-mix was comparable between the two periods. The analyses of financial productivity were based on Payment by Results (PbR) data for 2012–2013.

### 2.4.1 Bottom line results for the impact of surgical hubs on economic outcome measures

Evidence relating to the economic impact of surgical hubs is limited. There is evidence to suggest that surgical hubs may not be financially productive. However, this outcome was reported by a single study and as such firm conclusions cannot be made. There is evidence to suggest that total average costs are lower in surgical hubs when compared to general hospitals.

## 2.5 Impact of surgical hubs on patient reported outcomes

Five studies (three comparative and two non-comparative) assessed patient reported outcomes. These outcomes included quality of cataract care (patient satisfaction and perceived outcomes after surgery), Oxford Hip Score, patient satisfaction, and patient compliance. Both comparative and non-comparative study findings are described below.

### Quality of cataract care

One Dutch comparative study reported on quality of cataract care using quality indicators Net Promoter Score (NPS) and a patient-reported outcome measure (PROM) – both of which measure patient satisfaction and the perceived outcome of patients four weeks after cataract surgery respectively.

Kruse et al (2019a) reported mixed findings with respect to quality of care. ITCs scored significantly better on the NPS compared with general hospitals, while differences in the PROM scores were inconsistent and marginal. Based on PROM scores, ITCs seemed to perform better on patient satisfaction compared with general hospitals, but there were no differences in the patient-reported outcomes after cataract surgery.

### Oxford Hip Score

One UK comparative study reported on the Oxford Hip Score, which is a joint-specific, patient-reported outcome measure designed to assess disability in patients undergoing total hip replacement.

Joseph et al (2022) found that the Oxford Hip Score improvement at 6 weeks’ post-operative period was marginally higher in the pre-pandemic ward (18.8) compared to the protected elective surgical unit (16.4). This study did not control for pandemic-related factors.

### Patient satisfaction and compliance

Three studies (one comparative and two non-comparative studies) reported on patient satisfaction and compliance.

Kruse et al (2019b) aimed to explore a range of quality measures in Dutch independent treatment centres (ITC). Patient satisfaction ratings were based on location regarding treatment, information provision, listening competency, handling by staff, accommodation, and experience in scheduling an appointment. The study found that patients attending ITCs had an average satisfaction score of 8.74 ± 1.17. Chain membership (i.e., chain-affiliated ITCs) was found to have a negative influence on patient satisfaction.

Huddy et al (2022) reported the experience and patient outcomes from setting up a ‘COVID protected’ robotic unit for colorectal and renal surgery in a day-surgical unit attached to a hospital in UK. The study found that patient satisfaction on the ‘COVID protected’ unit was high, indicating that patients felt confident to undergo surgery at a time of increased risk.

Perrone et al (2020) described the experience of a Gynaecologic Oncology Unit’s reallocation of resources in a COVID-19 free surgical oncological hub to guarantee standard quality of surgical activities. This study explored patient’s compliance and satisfaction with the new COVID-19 free care setting and found that patients were compliant and well accepted the lifestyle restrictions and reorganisation of the care.

### 2.5.1 Bottom line results for the impact of surgical hubs on patient reported outcome measures

The evidence relating to the impact of surgical hubs on patient reported outcomes is limited but indicates there may be a positive effect on patient satisfaction and compliance. However, this evidence originates mostly from non-comparative studies.

## 3. DISCUSSION

### 3.1 Summary of the findings

There is evidence to suggest that surgical hubs can be effective at improving a range of clinical, performance, and patient reported outcomes in patients undergoing certain types of surgery. However, the evidence relating to the efficiency and financial productivity of these hubs is inconsistent. Considerable variation in the types of surgical hubs reviewed, surgical disciplines, along with the small number of comparative studies, as well as methodological limitations across included studies, could limit the applicability of these findings. It is worth noting that some of the outcomes reported in this review are likely to have been impacted by the COVID-19 pandemic. More urgent cases with higher rates of complexities and difficulties, were more likely to get referred for surgery during this period. These could account for findings such as the high reoperation rates reported during the peak phase of the pandemic, as well as quicker than usual discharge from hospital.

### 3.2 Limitations of the available evidence

This rapid review has highlighted several evidence gaps and areas of uncertainty. These include:

A small number of comparative studies were included in this review, and evidence for many outcomes were derived from single studies.

There appears to be a paucity of robust study designs in this area of research. The majority of included studies utilised weak research methodologies that may not be appropriate for inferring effectiveness. Some studies may be at risk of selection bias as details on patient characteristics are lacking. The methodological reporting in the included studies was also often inadequate, preventing a fully informed assessment of quality.

Included studies were focussed on various surgical specialties – some studies included a mix of surgical specialities. It is unclear whether the evidence derived from this review can be applied to other surgical fields.

Key details pertaining to information about the structure of surgical hubs were often lacking or poorly described in the included studies. Only two studies explicitly described surgical facilities with ring-fenced or dedicated capacity/resources.

The surgical hubs identified in this rapid review were varied in structure, function, and location. This could limit the generalisability of our findings.

The majority of included studies described surgical hubs that were established during the COVID-19 pandemic and designed mainly to mitigate the spread of COVID-19. This review may therefore not be able to address issues such as those pertaining to the impact of emergency/winter pressures.

Included studies that were conducted during the COVID-19 pandemic did not appear to control for the impact of the pandemic on outcomes such as length of hospital stay and reoperation rates.

This review initially sought to address a focussed review question and several sub-questions. However, no evidence was identified for most sub-questions. Furthermore, this review did not identify any evidence relating to resource use, safety outcomes, or willingness to travel (staff/patient) to surgical hub facilities.

### 3.3 Implications for policy and practice

This report has provided insights into how surgical hubs deliver services in distinct surgical areas. This may be useful when designing research or services to assist with the recovery of planned care services in the UK. Considering the paucity of robust evidence, further well-designed, higher quality research from the UK and similar countries is needed to better understand the effectiveness of surgical hubs in Wales.

### 3.4 Strengths and limitations of this Rapid Review

The studies included in this rapid review were identified through an extensive search of electronic databases, grey literature and the use of supplementary methods. Despite making every effort to capture all relevant publications and reduce the risk of bias in our review process, it is possible that additional eligible publications may have been missed and that biases do exist in this review.

To ensure the usefulness of our findings, only comparative studies were analysed when evaluating effectiveness outcomes, as these are better placed to determine the presence of cause-and-effect relationships. However, data from both comparative and non-comparative studies were used to provide insight into patient-reported outcomes such as patient satisfaction and compliance.

Due to the variability and paucity of evidence for particular outcomes, we did not attempt to undertake any assessment of the outcomes using GRADE. However, given the paucity of the evidence and the methodological limitations inherent in our included studies, caution needs to be applied when interpreting the findings of this review.

## Data Availability

All data produced in the present study are available upon reasonable request to the authors

## Abbreviations

Acronym: Full Description
ACL: Anterior Cruciate Ligament
AHT: Acute Hospital Trust
ASA: American Society of Anaesthesiologists
BPH: BUPA Cromwell Hospital
CND: Cataract National Dataset
CI: onfidence Interval
COVID-19: Coronavirus Disease 2019
CPG: Clinical Prioritisation Group
CRS: Colorectal Surgery
CTS: Cataract, and Carpal Tunnel Surgeries
DRG: Diagnostic-Related Group System
ERP: Enhanced Recovery Protocols
GO: Gynaecological Oncology
ISTC: Independent Sector Treatment Centres
ITC: Independent Treatment Centres
JBI: Joanna Briggs Institute
MRSA: Methicillin-Resistant Staphylococcus Aureus
NHS: National Health Service
NPS: Net Promoter Score
OR: Odds Ratio
OSH: Oncological Surgical Hub
PbR: Payment by Results
PESU: Protected Elective Surgical Units
PPW: Pre-Pandemic Ward
PROM: Patient Reported Outcome Measures
RMH: Royal Marsden Hospital
RR: Rapid Review
SARS-CoV-2: Severe Acute Respiratory Syndrome Coronavirus 2
THR: Total Hip Replacement
TKR: Total Knee Replacement
UK: United Kingdom
UKSH: UK Specialist Hospitals

## 5. RAPID REVIEW METHODS

### 5.1 Eligibility criteria

We searched for primary sources to answer the review question: “what is the effectiveness, efficiency, and acceptability of surgical hubs in supporting planned care activity, and how best should they be established and run?” The following eligibility criteria were used to identify studies for inclusion in the rapid review:

**Table 2:** Eligibility criteria

|  | Inclusion criteria | Exclusion criteria |
| --- | --- | --- |
| Population | Surgical patients of all ages (children and adults)<br>All elective surgical fields | Emergency or transplant surgery |
| Intervention / exposure | Elective surgical hubs defined as protected facilities dedicated entirely to elective care, with ring-fenced resources that allow them to stay active even when emergency pressures rise (Briggs et al., 2022)<br>To include stand-alone, integrated, and ring-fenced surgical hubs. Can be privately or NHS-run facilities or mixed models<br>Surgical hubs can include day cases, one or more specialties, short stay units with or without enhanced recovery | Facilities dedicated to non-elective surgical care<br>Outpatient surgical facilities e.g., ambulatory surgery centres, outpatient surgery centres<br>Referral to an alternative surgical site, which does not meet the definition of a surgical hub, for example due to an excessive waiting list |
| Comparison | No comparison<br>Usual care<br>Other service/governance or workforce models<br>Other time period or non-emergency situation |  |
| Outcome measures | Studies reporting any of the following outcomes will be considered for inclusion in the review:<br>• Clinical outcomes<br>• Performance outcomes<br>• Economic outcomes<br>• Safety outcomes<br>• Resource use<br>• Patient experience/attitudes<br>• Acceptability (patients/staff)<br>• Willingness to travel |  |
| Study design | Primary studies: comparative and non-comparative quantitative studies or qualitative studies, full economic evaluations | Secondary/tertiary research<br>Commentaries, Editorials |
| Countries | High-income countries |  |
| Language of publication | English |  |
| Publication type | Published, preprint |  |
| Other factors<br>*Any other key points to note* | Papers published since 2015 |  |

### 5.2 Literature search

The studies included in this rapid review were identified through a systematic literature search. General repositories of evidence noted in our resource list were searched between the 13^th^ and 17^th^ of January 2023. Grey literature searching and citation tracking of the secondary sources included in scoping work were also conducted to identify any additional studies. An audit trail of the search process is provided within the resource list (Appendix 1). Searches were limited to English-language publications that were published since 2015 and included searches for primary studies.

Search concepts and keywords around surgical hubs/ centres and elective surgery were utilised. The searches included free text words and we deliberately kept our search strategy broad to capture as much evidence on surgical hubs as possible. The search strategy used to search MEDLINE is available in Appendix 2.

### 5.3 Study selection process

The 513 studies identified through the database and grey literature searches were uploaded to the systematic reviewing platform Rayyan for title and abstract screening. Two independent reviewers screened title and abstracts against the eligibility criteria in Table 2, with disagreements resolved by a third reviewer. Sixty-two articles were screened at full text by two independent reviewers with disagreements resolved by a third reviewer. In total, 12 studies were included following full text screening. A visual representation of the flow of studies throughout the review can be found in Figure 2.

### 5.4 Data extraction

One researcher performed the data extraction and a second researcher carried out consistency checks. Information extracted includes:

- Reference (author, year, country)
- Study design
- Intervention / comparator
- Aim
- Data collection methods (and dates)
- Outcome(s) measured
- Study participants (e.g., sample size)
- Setting
- Surgical speciality
- Key findings
- Additional notes/comments to report key information not captured in the above, and to record any limitations of the included sources.

### 5.5 Quality appraisal

A range of JBI quality appraisal checklists (which were selected based on the study design used) were used to assess the methodological quality of each included study. Quality assessment was undertaken by a single reviewer, with verification of all judgements by a second reviewer. Any discrepancies were discussed and resolved amongst the review team. The results of the quality appraisals can be seen in Tables 3, 4, and 5.

### 5.6 Synthesis

A narrative synthesis was conducted reporting results from included studies with a comparative element. A map outlining outcomes assessed in the included studies was created in order to visualise the breadth of the evidence identified and the evidence gaps (Figure 1).

## 6. EVIDENCE

### 6.1 Study selection

**Figure 2.**
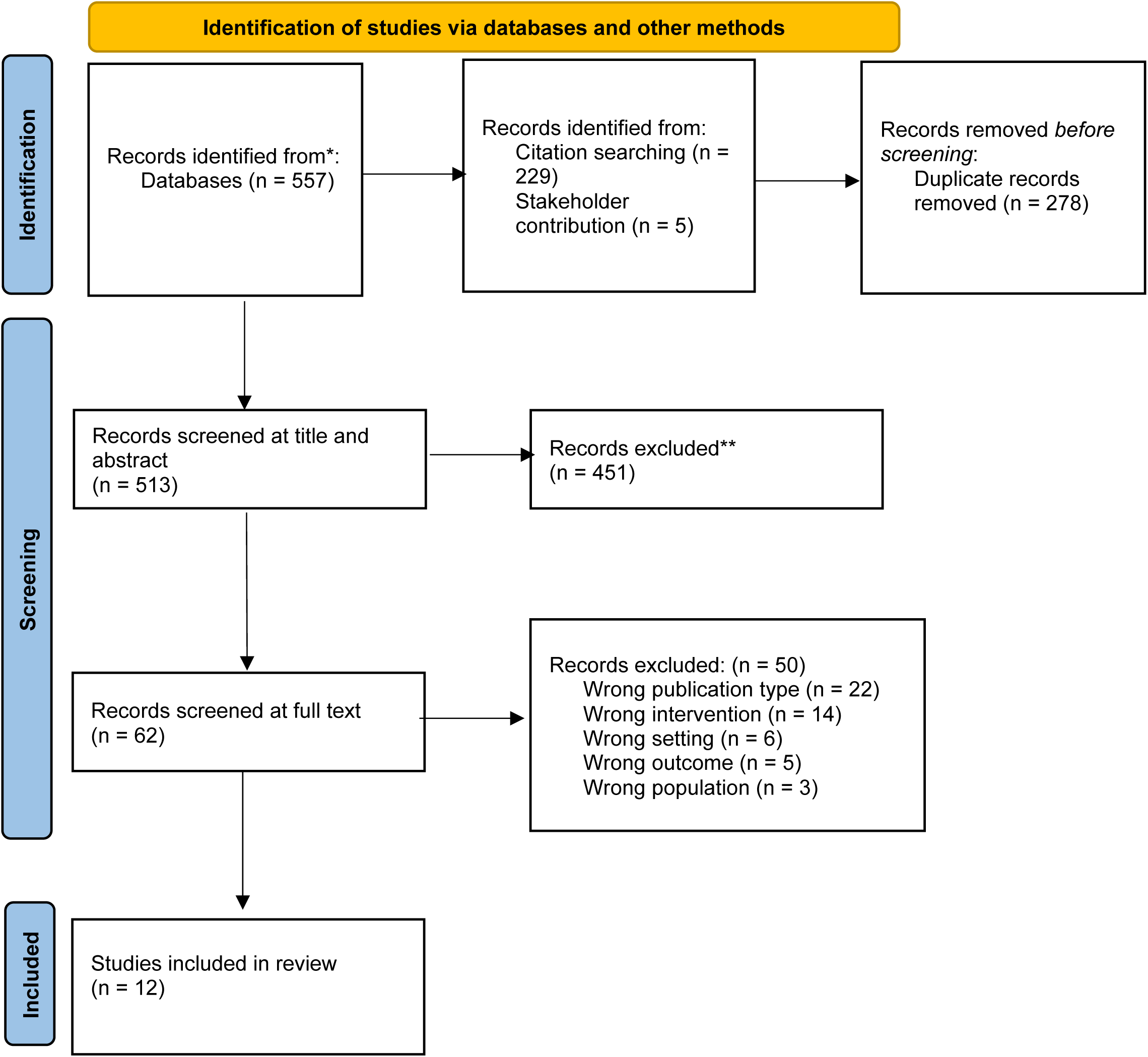
PRISMA flow chart

### 6.2 Quality appraisal tables

**Table 3.** Quality appraisal results for quasi-experimental studies

| Study | JBI Appraisal Items – Quasi-experimental studies |  |  |  |  |  |  |  |  |
| --- | --- | --- | --- | --- | --- | --- | --- | --- | --- |
|  | Q1. | Q2. | Q3. | Q4. | Q5. | Q6. | Q7. | Q8. | Q9. |
| Joseph et al 2022 | Y | Y | N | Y | N | N | Y | Y | Y |
| Kruse et al 2019a | Y | N | N | Y | N/A | N/A | Y | Y | Y |
| Spinelli et al 2021 | Y | Y | Y | Y | N/A | Y | Y | Y | Y |
| Syed et al 2015 | Y | U | U | Y | N | Y | N | Y | Y |
| Tulp et al 2020 | Y | U | Y | Y | N | N | Y | Y | Y |
| Vanhegan et al 2015 | Y | U | Y | Y | N | N/A | Y | Y | Y |
| Key: Y=Yes, N=No, U=Unclear, N/A=not applicable |  |  |  |  |  |  |  |  |  |

1. Is it clear what is the cause and what is the effect? 2. Were the participants included in any comparisons similar? 3. Were the participants included in any comparisons receiving similar treatment/care, other than the exposure or intervention of interest? 4. Was there a control group? 5. Were there multiple measurements of the outcome both pre and post the intervention/ exposure? 6. Was follow up complete and if not, were differences between groups in terms of their follow up adequately described and analysed? 7. Were the outcomes of participants included in any comparisons measured in the same way? 8. Were outcomes measured in a reliable way? 9. Was appropriate statistical analysis used?

**Table 4.**
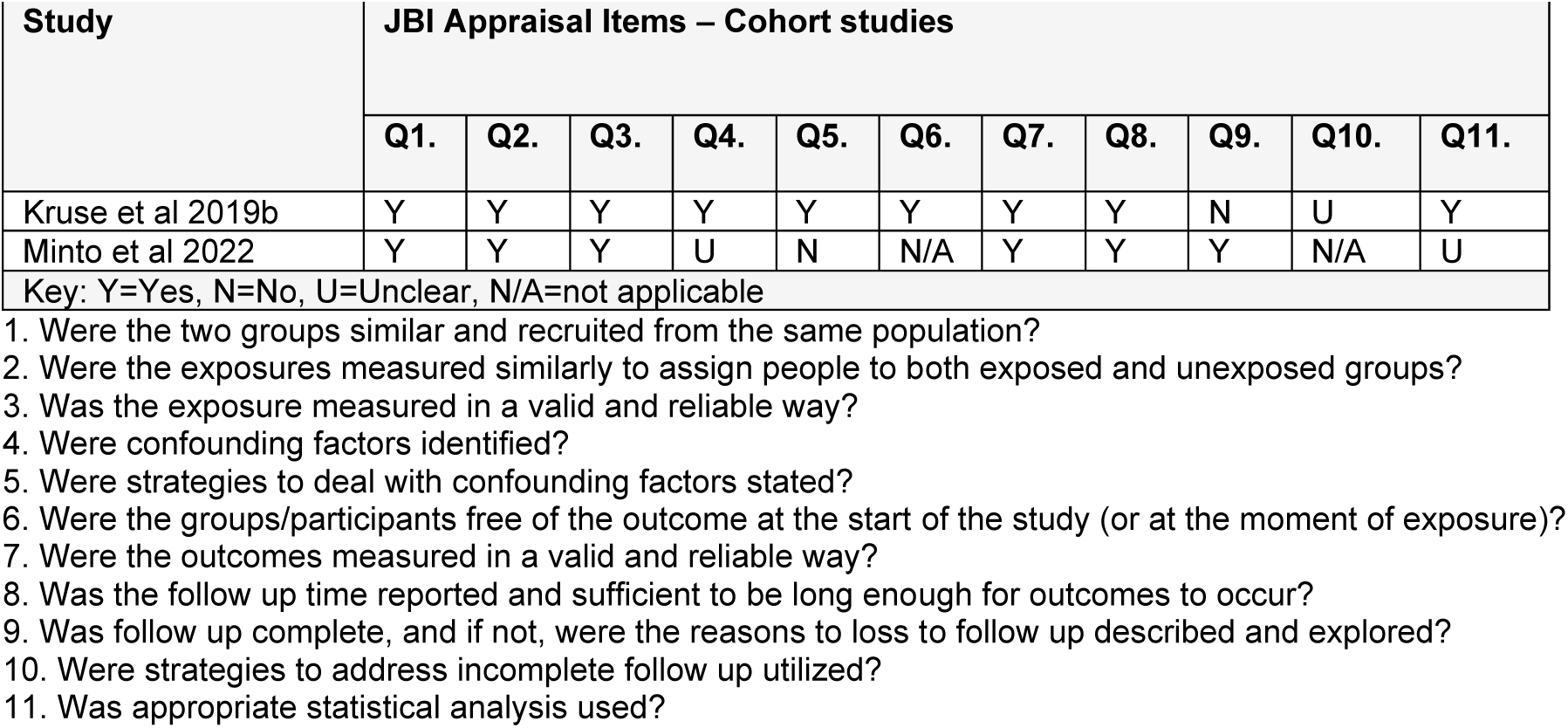
Quality appraisal results for Cohort studies

**Table 5.** Quality appraisal results for case series studies

| Study | JBI Appraisal Items – Case series studies |  |  |  |  |  |  |  |  |  |
| --- | --- | --- | --- | --- | --- | --- | --- | --- | --- | --- |
|  | Q1. | Q2. | Q3. | Q4. | Q5. | Q6. | Q7. | Q8. | Q9. | Q10. |
| Carvalho et al 2022 | Y | Y | Y | N | N | Y | Y | Y | Y | Y |
| Huddy et al 2022 | Y | Y | Y | N | U | Y | N | Y | Y | Y |
| Jeanon et al 2021 | Y | Y | Y | Y | N | N | Y | Y | Y | Y |
| Perrone et al 2021 | Y | Y | Y | N | N | Y | Y | N | Y | Y |
| Key: Y=Yes, N=No, U=Unclear, N/A=not applicable |  |  |  |  |  |  |  |  |  |  |

1. Were there clear criteria for inclusion in the case series? 2. Was the condition measured in a standard, reliable way for all participants included in the case series? 3. Were valid methods used for identification of the condition for all participants included in the case series? 4. Did the case series have consecutive inclusion of participants? 5. Did the case series have complete inclusion of participants? 6. Was there clear reporting of the demographics of the participants in the study? 7. Was there clear reporting of clinical information of the participants? 8. Were the outcomes or follow up results of cases clearly reported? 9. Was there clear reporting of the presenting site(s)/clinic(s) demographic information? 10. Was statistical analysis appropriate?

## 7. ADDITIONAL INFORMATION

### 7.2 Acknowledgements

The Public Health Wales team would like to thank Lesley Law, Luke John Davies, and Alexandra Strong for their time expertise and contributions during stakeholder meetings in guiding the focus of the review and interpretation of findings.

### 7.3 Disclaimer

The views expressed in this publication are those of the authors, not necessarily Health and Care Research Wales. The Health and Care Research Wales Evidence Centre and authors of this work declare that they have no conflict of interest.

## 8. ABOUT THE HEALTH AND CARE RESEARCH WALES EVIDENCE CENTRE

The Health and Care Research Wales Evidence Centre integrates with worldwide efforts to synthesise and mobilise knowledge from research.

We operate with a core team as part of <u>Health and Care Research Wales</u>, Welsh Government and are led by Professor Adrian Edwards of Cardiff University.

The core team of the centre works closely with collaborating partners in the Bangor Institute <u>for Health and Medical Research (BIHMR)</u>, Bangor University, which includes the <u>Centre for</u> <u>Health Economics and Medicines Evaluation (CHEME)</u> working in collaboration with Health and Care Economics Cymru, <u>Health Technology Wales</u>, <u>Public Health Wales Evidence</u> <u>Service</u>, <u>Population Data Science, Swansea University</u> using <u>SAIL Databank</u>, the <u>Wales</u> <u>Centre for Evidence Based Care (WCEBC)</u>, the <u>Specialist Unit for Review Evidence (SURE)</u> and <u>CASCADE</u>, Cardiff University.

**Director**: Professor Adrian Edwards

**Website**: www.researchwalesevidencecentre.co.uk

## APPENDIX

### APPENDIX 1: Resources searched during Rapid Review Searching

**Table 6:**
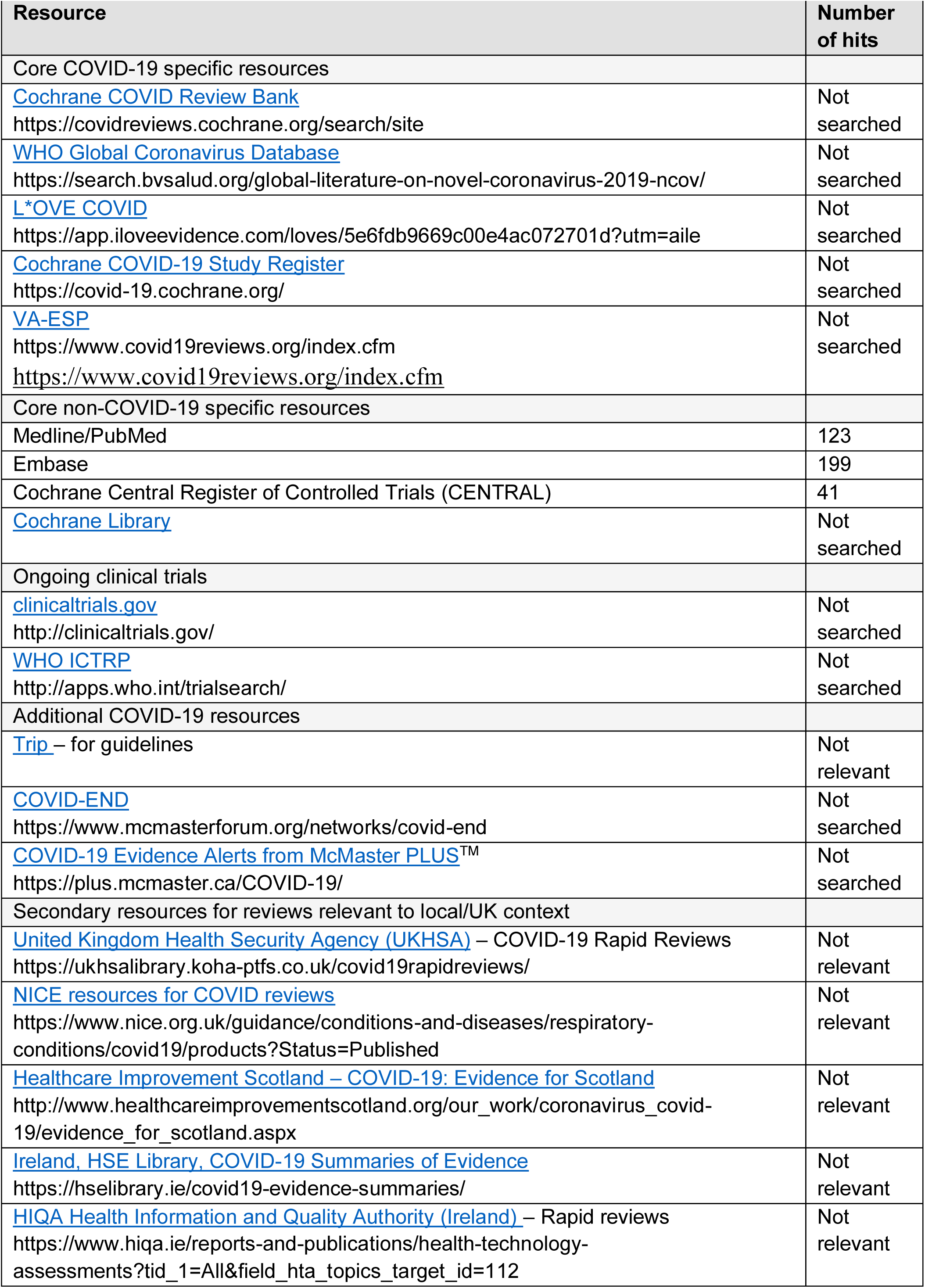
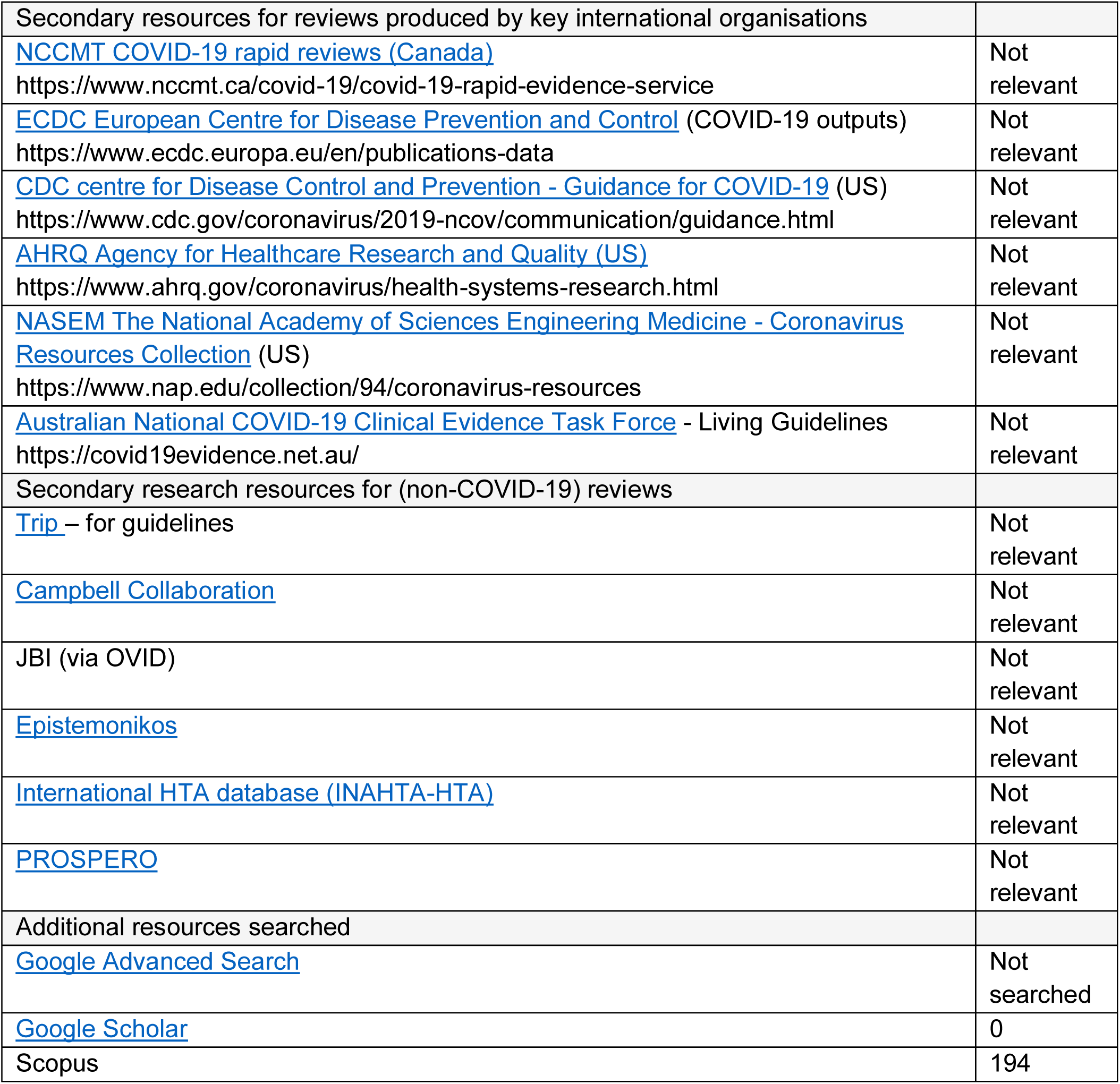
Resources searched

| Resource | Number of hits |
| --- | --- |
| Core COVID-19 specific resources |  |
| [Cochrane COVID Review Bank](https://covidreviews.cochrane.org/search/site)<br>https://covidreviews.cochrane.org/search/site | Not searched |
| [WHO Global Coronavirus Database](https://search.bvsalud.org/global-literature-on-novel-coronavirus-2019-ncov/)<br>https://search.bvsalud.org/global-literature-on-novel-coronavirus-2019-ncov/ | Not searched |
| [L*OVE COVID](https://app.iloveevidence.com/loves/5e6fdb9669c00e4ac072701d?utm=aile)<br>https://app.iloveevidence.com/loves/5e6fdb9669c00e4ac072701d?utm=aile | Not searched |
| [Cochrane COVID-19 Study Register](https://covid-19.cochrane.org/)<br>https://covid-19.cochrane.org/ | Not searched |
| [VA-ESP](https://www.covid19reviews.org/index.cfm)<br>https://www.covid19reviews.org/index.cfm<br>https://www.covid19reviews.org/index.cfm | Not searched |
| Core non-COVID-19 specific resources |  |
| Medline/PubMed | 123 |
| Embase | 199 |
| Cochrane Central Register of Controlled Trials (CENTRAL) | 41 |
| Cochrane Library | Not searched |
| Ongoing clinical trials |  |
| clinicaltrials.gov<br>http://clinicaltrials.gov/ | Not searched |
| [WHO ICTRP](http://apps.who.int/trialsearch/)<br>http://apps.who.int/trialsearch/ | Not searched |
| Additional COVID-19 resources |  |
| Trip – for guidelines | Not relevant |
| [COVID-END](https://www.mcmasterforum.org/networks/covid-end)<br>https://www.mcmasterforum.org/networks/covid-end | Not searched |
| [COVID-19 Evidence Alerts from McMaster PLUS™](https://plus.mcmaster.ca/COVID-19/)<br>https://plus.mcmaster.ca/COVID-19/ | Not searched |
| Secondary resources for reviews relevant to local/UK context |  |
| [United Kingdom Health Security Agency (UKHSA) – COVID-19 Rapid Reviews](https://ukhsalibrary.koha-ptfs.co.uk/covid19rapidreviews/)<br>https://ukhsalibrary.koha-ptfs.co.uk/covid19rapidreviews/ | Not relevant |
| [NICE resources for COVID reviews](https://www.nice.org.uk/guidance/conditions-and-diseases/respiratory-conditions/covid19/products?Status=Published)<br>https://www.nice.org.uk/guidance/conditions-and-diseases/respiratory-conditions/covid19/products?Status=Published | Not relevant |
| [Healthcare Improvement Scotland – COVID-19: Evidence for Scotland](http://www.healthcareimprovementscotland.org/our_work/coronavirus_covid-19/evidence_for_scotland.aspx)<br>http://www.healthcareimprovementscotland.org/our_work/coronavirus_covid-19/evidence_for_scotland.aspx | Not relevant |
| [Ireland, HSE Library, COVID-19 Summaries of Evidence](https://hselibrary.ie/covid19-evidence-summaries/)<br>https://hselibrary.ie/covid19-evidence-summaries/ | Not relevant |
| [HIQA Health Information and Quality Authority (Ireland) – Rapid reviews](https://www.hiqa.ie/reports-and-publications/health-technology-assessments?tid_1=All&field_hta_topics_target_id=112)<br>https://www.hiqa.ie/reports-and-publications/health-technology-assessments?tid_1=All&field_hta_topics_target_id=112 | Not relevant |
| Secondary resources for reviews produced by key international organisations |  |
| [NCCMT COVID-19 rapid reviews (Canada)](https://www.nccmt.ca/covid-19/covid-19-rapid-evidence-service)<br>https://www.nccmt.ca/covid-19/covid-19-rapid-evidence-service | Not relevant |
| [ECDC European Centre for Disease Prevention and Control](https://www.ecdc.europa.eu/en/publications-data) (COVID-19 outputs)<br>https://www.ecdc.europa.eu/en/publications-data | Not relevant |
| [CDC centre for Disease Control and Prevention - Guidance for COVID-19](https://www.cdc.gov/coronavirus/2019-ncov/communication/guidance.html) (US)<br>https://www.cdc.gov/coronavirus/2019-ncov/communication/guidance.html | Not relevant |
| [AHRQ Agency for Healthcare Research and Quality (US)](https://www.ahrq.gov/coronavirus/health-systems-research.html)<br>https://www.ahrq.gov/coronavirus/health-systems-research.html | Not relevant |
| [NASEM The National Academy of Sciences Engineering Medicine - Coronavirus Resources Collection](https://www.nap.edu/collection/94/coronavirus-resources) (US)<br>https://www.nap.edu/collection/94/coronavirus-resources | Not relevant |
| [Australian National COVID-19 Clinical Evidence Task Force](https://covid19evidence.net.au/) - Living Guidelines<br>https://covid19evidence.net.au/ | Not relevant |
| Secondary research resources for (non-COVID-19) reviews |  |
| Trip – for guidelines | Not relevant |
| Campbell Collaboration | Not relevant |
| JBIC (via OVID) | Not relevant |
| Epistemonikos | Not relevant |
| International HTA database (INAHTA-HTA) | Not relevant |
| PROSPERO | Not relevant |
| Additional resources searched |  |
| Google Advanced Search | Not searched |
| Google Scholar | 0 |
| Scopus | 194 |

### APPENDIX 2: Search strategy used for MEDLINE

| Set# | Searched for | Results |
| --- | --- | --- |
| 1 | (green pathway and surg*).tw. | 4 |
| 2 | (surg* adj2 hub*).tw. | 32 |
| 3 | (elective adj2 surg* adj2 (centre* or center* or clinic or clinics or unit or units)).tw. | 101 |
| 4 | (elective adj2 orthop?edic* adj2 (centre* or center* or clinic or clinics or unit or units)).tw. | 34 |
| 5 | (elective centre* or elective center* or elective care centre hub*).tw. | 6 |
| 6 | "out of hospital surgical facilit*".tw. | 0 |
| 7 | chartered surg* facilit*.tw. | 0 |
| 8 | "non hospital surg* facilit*".tw. | 1 |
| 9 | hospital associated surg* cent*.tw. | 0 |
| 10 | (independent adj2 treatment adj (center* or centre*)).tw. | 47 |
| 11 | minor surg* setting*.tw. | 3 |
| 12 | or/1-11 | 220 |
| 13 | limit 12 to (english language and yr="2015 -Current") | 123 |

## REFERENCES

British Medical Association (2023). NHS backlog data analysis. Available at: https://www.bma.org.uk/advice-and-support/nhs-delivery-and-workforce/pressures/nhs-backlog-data-analysis [Accessed 13 March 2023].

Briggs T, Kay P, Vig S, et al. (2022). Optimising surgical hubs for staff: case studies on training, wellbeing and retention. British Journal of Healthcare Management, 1–9.

Department of Health and Social Care (2022). Over 50 new surgical hubs set to open across England to help bust the COVID-19 backlogs. Available at: https://www.gov.uk/government/news/over-50-new-surgical-hubs-set-to-open-across-england-to-help-bust-the-covid-backlogs. [Accessed 10 December 2022].

GIRFT (2022). Design and layout of elective surgical hubs - a guide for NHS systems and regions to support planning of effective surgical hubs. NHS.

Huddy J R, Crockett M, Nizar A S, et al. (2022). Experiences of a “COVID protected” robotic surgical centre for colorectal and urological cancer in the COVID-19 pandemic. Journal of Robotic Surgery, 16, 59–64.

Joseph V, Boktor J G E, Roy K, et al. (2022). Dedicated orthopaedic elective unit: our experience from a district general hospital. Journal of Medical Science, 1971-.

Mallorie, S. (2023). What caused the UK’s elective care backlog, and how can we tackle it? The King’s Fund. Available at: https://www.kingsfund.org.uk/blog/2023/02/what-caused-uks-elective-care-backlog-how-can-we-tackle-it [Accessed 13 March 2023].

Minto T, Abdelrahman T, Jones L, et al. (2022). Safety of maintaining elective and emergency surgery during the COVID-19 pandemic with the introduction of a Protected Elective Surgical Unit (PESU): A cross-specialty evaluation of 30-day outcomes in 9,925 patients undergoing surgery in a University Health Board. Surg Open Sci, 10, 168–173.

Royal College of Surgeons of England (2022). The case for surgical hubs. London: The Royal College of Surgeons of England. Available at: https://www.rcseng.ac.uk/about-the-rcs/government-relations-and-consultation/position-statements-and-reports/the-case-for-surgical-hubs/

Welsh Government (2022). Our programme for transforming and modernising planned care and reducing waiting lists in Wales. Available at: https://www.gov.wales/transforming-and-modernising-planned-care-and-reducing-nhs-waiting-lists

